# Population-scale genomics reveals divergent pathogenicity of variant classes across paralogous collagen IV genes

**DOI:** 10.64898/2026.06.12.26355529

**Authors:** Konstantinos Tzoumkas, Gabriel T. Doctor, Omid Sadeghi-Alavijeh, Daniel P. Gale

## Abstract

Monoallelic pathogenic or likely pathogenic variants in *COL4A3* and *COL4A4* occur in approximately 1 in 106 individuals, yet whether these paralogous genes confer equivalent pathogenicity for the same variant classes has not been tested at population scale. Using whole-genome sequencing data from the UK Biobank (UKB; n ≈ 500,000), with replication in the All of Us Research Program (n ≈ 414,000), we performed per-variant association testing, gene-based collapsing analyses and phenome-wide association studies (PheWAS) across haematuria, proteinuria and chronic kidney disease. We identified 64 *COL4A3* and 92 *COL4A4* rare variants significantly associated with haematuria or proteinuria, generating a quantitative allelic series for clinical variant interpretation. Glycine substitutions within collagenous domains conferred similar risks in both genes. In contrast, truncating and non-collagenous domain (NC1) missense variants were strongly associated with haematuria and proteinuria in *COL4A4* carriers but showed substantially attenuated or absent associations in *COL4A3* carriers despite comparable carrier frequencies and predicted pathogenicity scores. These findings were independently replicated in All of Us. Genome-wide association analysis identified the *COL4A3/COL4A4* locus as the dominant genetic determinant of haematuria, with the signal attributable to the aggregate effects of rare coding variants and no evidence of independent common variant or trans-acting modifier effects. These findings demonstrate substantial gene-specific differences in tolerance to truncating and NC1 variants between *COL4A3* and *COL4A4*, challenging assumptions of equivalent pathogenicity across paralogous collagen IV genes. Gene identity and not variant class alone, should inform risk stratification, variant interpretation and genetic counselling in individuals carrying collagen IV risk genotypes.

## Introduction

Alport Syndrome (AS) is a hereditary kidney disorder caused by pathogenic variants in the paralogous genes *COL4A3*, *COL4A4*, and *COL4A5*, which respectively encode the Collagen α3(IV), α4(IV), and α5(IV) chains that are made by glomerular podocytes and trimerize to form the Type IV Collagen that is essential for glomerular basement membrane (GBM) and podocyte integrity and function^1^. Pathogenic variants in these genes can disrupt the glomerular filtration barrier, leading to haematuria (typically first noted during childhood) with subsequent podocyte loss, proteinuria and progressive kidney damage resulting in chronic kidney disease (CKD) and kidney failure (KF) which can occur as early as childhood in some individuals and as late as old age in others. The syndrome is also associated with sensorineural hearing loss (which tends to mirror kidney disease in its severity) and ocular abnormalities^2^.

X-linked AS (XLAS), caused by pathogenic *COL4A5* variants, is most severe in hemizygous males in whom there is a high lifetime risk of kidney failure, while heterozygous females usually exhibit milder disease, although can manifest CKD and indeed KF^3^. Autosomal recessive AS (ARAS), due to biallelic pathogenic *COL4A3* or *COL4A4* variants, affects both sexes similarly and typically causes early-onset, severe kidney disease^4^. Heterozygous pathogenic variants in the autosomal *COL4A3* or *COL4A4* genes are strongly associated with microscopic haematuria with (where kidney biopsy is performed) thinning of the GBM, and are now recognized to be risk factors for proteinuria and even KF, which is thought to have a lifetime risk among heterozygotes of up to 3%.^5^

Type IV Collagen includes a triple helical collagenous domain composed of a repeating Gly-X-Y motif interspersed with non-collagenous interruptions, and a C-terminal non-collagenous (NC1) domain crucial to initiate trimer formation^6^. Glycine, as the smallest amino acid, is necessary for tight winding of the triple helix and its substitution with a more bulky amino acid is the commonest type of mutation in AS. Pathogenic missense variants that affect NC1 domain can have a stronger loss-of-function effect since they can abrogate formation of the trimer and consequently, protein truncating *COL4A5* variants in hemizygous males, and biallelic truncating *COL4A3* or *COL4A4* variants in either sex, are associated with younger age at kidney failure than are Glycine substitutions. Disease risk and genotype-phenotype correlations are less well understood in individuals with heterozygous autosomal variation with some data suggesting a lower KF risk for truncating variants^7,8^ and much of the literature in this group biased by the effects of ascertainment (owing to the fact that individuals with a personal or family history of kidney disease were more likely to have the relevant genes analysed than the healthy population)^9^. However, analysis of genomic data from cohorts not enriched for kidney disease suggest that up to 1 in 106 individuals carry a pathogenic *COL4A3*/4 variant in heterozygosity^4^, resulting in a combined frequency of such individuals great enough for population-level analyses to offer the possibility of better quantifying disease risk associated with heterozygous genotypes.

Clinical guidelines and variant classification frameworks have generally treated these genes as functionally interchangeable, but this assumption has not been tested at population scale and is potentially consequential for risk stratification. Here, we leverage UK Biobank genomic and clinical data to examine the associations between pathogenic *COL4A3*, *COL4A4*, and *COL4A5* variants and renal phenotypes, including haematuria, proteinuria, and CKD. By quantifying these relationships based on variant effect and zygosity, we aim to improve understanding of how the presence of a heterozygous variant can lead to disease and potentially identify genetic markers that can be used prognostically.

## Methods

### UK Biobank

The UK Biobank (UKB) is a large, ongoing prospective cohort study comprising approximately 500,000 mostly healthy adults from the United Kingdom, aged 40 to 69 years at the time of recruitment. It provides a comprehensive resource for population-based research by collecting health questionnaires, physical measurements, and biological samples, with ongoing linkage to electronic health records to track clinical outcomes over time^10^. All data, including centrally curated genetic and health information, are made available to approved researchers through an application process.

### Case/Control Definitions for Glomerular Phenotypes

Cases of glomerular haematuria were defined as individuals with one or more of the following ICD-10 codes from Hospital inpatient summary diagnoses and first occurrences: R31 (Unspecified haematuria), N02.8 (Other recurrent and persistent haematuria), or N02.9 (Recurrent and persistent haematuria, unspecified). To increase specificity for a glomerular aetiology and minimize misclassification, we excluded individuals with diagnostic codes indicative of major alternative causes of haematuria, including urinary tract infections, urolithiasis, and malignancies of the genitourinary tract (see Supplementary Table 1 for the complete list of exclusion codes). Participants without any haematuria codes and without any of the listed exclusion diagnoses were defined as controls.

Cases of glomerular proteinuria were defined using a composite approach to maximize sensitivity, incorporating both diagnostic codes and biomarker data. Individuals were considered cases if they had either the ICD-10 code R80 (Isolated proteinuria) recorded in hospital inpatient summary diagnostic data or a urinary albumin-to-creatinine ratio (uACR) > 3 mg/mmol, calculated from urinary albumin (Data-Field 30500) and urinary creatinine (Data-Field 30510) measured at baseline assessment. To increase specificity for a glomerular aetiology and maintain consistency with the haematuria definition, the same exclusion criteria were applied (Supplementary Table 1). Controls were defined as participants without the R80 code, with a uACR ≤ 3 mg/mmol, or without any of the listed exclusion diagnoses.

Cases of severe CKD were defined using a composite of diagnostic codes and procedure records. Individuals were classified as cases if they had one or more relevant ICD-10 codes for advanced CKD stages and end-stage renal disease, or relevant OPCS-4 procedure codes for kidney replacement therapy (see Supplementary Table 2 for the complete list of inclusion codes). Controls were defined as participants without any of the case-defining codes and without any codes listed in control exclusion criteria (see Supplementary Table 3 for the complete list of CKD control codes).

### *COL4A3/A4/A5* Per-variant analysis

Variants in *COL4A3*, *COL4A4* and *COL4A5* were extracted from the population-level whole-genome sequencing (WGS) data (DRAGEN pVCF format, UK Biobank 500k release v.20.5). To mitigate confounding by population stratification, the analysis was restricted to individuals of genetically inferred European ancestry. *COL4A5*, located on the X chromosome, was analysed separately for males and females to account for zygosity. For all variants within these genes, we performed a per-variant association test. Given the very low number of carriers for rare variants (1-3 individuals), odds ratios (ORs) and 95% confidence intervals (CIs) for association with haematuria, proteinuria and severe CKD were derived from 2×2 contingency tables using Fisher’s Exact Test. To ensure biological relevance and clinical interpretability, we focused subsequent reporting on variants that are coding and had statistical support (95% CI for the OR excluding 1) and further assessed their pathogenicity using computational predictions such as CADD v.1.7^11^ and REVEL^12^.

### *COL4A3/A4* Collapsing and Phenome-Wide Association Study (PheWAS)

To investigate the aggregate phenotypic burden of rare *COL4A3* and *COL4A4* variants, we performed gene-based collapsing analyses followed by phenome-wide association study (PheWAS). Rare (MAF < 1%) predicted pathogenic variants in each gene were classified into 3 mutually exclusive functional categories per gene: Glycine (Gly) substitutions: missense variants affecting the conserved glycine residue within the Gly-X-Y repeat motif of the collagenous domain with a CADD score > 20; Truncating variants (Trunc): predicted high-confidence (HC) loss-of-function (lof) variants (nonsense, frameshift, etc) as annotated by LOFTEE (v.1.0.4)^13^; and NC1 domain missense variants (NC1): missense variants located specifically within the C-terminal non-collagenous (NC1) domain with a CADD score > 20. For each gene, variants within a class were collapsed into a binary carrier status, creating 6 distinct genetic predictors.

The PheWAS was conducted against a panel of 1,363 unique binary ICD10 phenotypes derived from the UK Biobank hospital inpatient records (Summary Diagnosis; First Occurrences). The pre-specified traits of the proteinuria (uACR > 3 mg/mmol), and severe CKD (stages 4-5/ESRD) were integrated into this phenotype list. For each phenotype, analyses were restricted to individuals without any diagnoses in the corresponding ICD-10 chapter. All collapsing tests were performed using SAIGE (Scalable and Accurate Implementation of Generalized mixed model v.1.5.0.2)^14^, adjusting for age, sex, and the first 10 genetic principal components, which generated both Burden and SKAT-O p-value for each variant class-phenotype association.

### Replication in the All of Us Cohort

To validate our primary findings, we replicated the collapsing PheWAS analyses in the All of Us (AoU) Research Program Curated Data Repository (CDR) version 8 (CDRv8)^15,16^. We restricted the AoU cohort to participants of European ancestry aged ≥40 years to match the demographic profile of the UK Biobank. The same 6 binary genetic burden scores (Gly-sub, Truncating and NC1 variant classes for *COL4A3* and *COL4A4*) were constructed using identical variant classification criteria. PheWAS was performed against 1,322 unique ICD-10 phenotypes derived from the AoU electronic health records, using SAIGE (v.1.5.0.2) with the same adjustments and same statistical models. Phenotype definitions and case/control criteria were applied consistently. For proteinuria, which could not be defined using a unified laboratory measurement across the cohort, we identified individuals with relevant diagnostic codes, urine dipstick measurements, or abnormal albumin/creatinine concentrations from the laboratory measurements data, applying a threshold of uACR > 3 mg/mmol or equivalent where available. This replication analysis served as an independent confirmation of the associations identified in the UK Biobank.

### Comparison of Genetic Effect Sizes

To formally test differences in the phenotypic impact of variant classes between *COL4A3* and *COL4A4*, we compared the estimated effect sizes (BETAs) and their standard errors (SEs) from the SAIGE PheWAS models for haematuria and proteinuria. This comparison was performed separately for each variant class (Gly-sub, Truncating, NC1) and for both UK Biobank and All of Us cohorts. For a given trait and variant class, we calculated the difference in BETA estimates between *COL4A3* and *COL4A4*

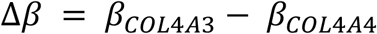

with its corresponding standard error

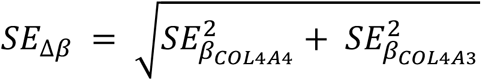

and the significance of this difference was assessed using two-tailed Z-test

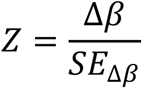

The validity of this test relies on the asymptomatic normality of the maximum likelihood coefficient estimates, an assumption well-supported by the substantial number of variant carriers underlying each burden test. A significant result (p < 0.05) indicated a statistically different genetic effect between the two genes for that specific variant class and phenotype.

### Genome-Wide Association Study for Glomerular Haematuria

A genome-wide association study (GWAS) was performed to identify genetic loci associated with glomerular haematuria, using the previously defined cases/control phenotype. To minimize confounding due to population structure and relatedness, we filtered for unrelated individuals using the KING software removing 3^rd^ degree relatives or closer. From this unrelated set, controls were ancestry-matched to cases at a 1:5 ratio using genetic principal components. The GWAS was conducted using SAIGE (v.1.5.0.2) on the machine learning (ML)-corrected, population-level WGS variant data in PLINK2 format (UK biobank 500k release v.20.5). The model was adjusted for age, sex and the first 10 genetic principal components.

To identify secondary genetic modifiers acting independently of the primary *COL4A3/A4* signal, we performed a second GWAS for glomerular haematuria which used a weighted *COL4A3/A4* burden score (WBS) as a continuous covariate. The WBS was calculated per individual as

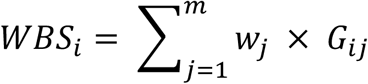

where *G_ij_* the genotype dosage for individual *i* at rare variant *j* in *COL4A3* or *COL4A4* and *w_j_* is the variant-specific weight, defined as the per-variant OR for glomerular haematuria (95% CI excluding 1) were included in the WBS. This adjusted GWAS was performed using the same SAIGE framework and covariates as the unadjusted GWAS with the addition of the WBS.

### GWAS Restricted to *COL4A3/A4* Heterozygotes

We performed a secondary GWAS restricted to heterozygotes for any qualifying *COL4A3* or *COL4A4* variant to investigate genetic modifiers operating specifically in these individuals. Qualifying variants were defined as glycine substitutions, truncating variants and NC1 domain missense variants with CADD score > 20. The analysis was performed using the same methodology as the primary GWAS, including filtering for unrelated individuals and adjustment for age, sex and the first 10 genetic principal components using SAIGE (v.1.5.0.2). This GWAS was conducted both without and with adjustment for the WBS as continuous covariate, as described above.

## Results

### Per-Variant Association with Glomerular Haematuria

A total of 9,161 haematuria cases (50.8% male) and 335,746 controls (42.6% male) of European ancestry were included in the analysis. A total of 131 rare coding variants were statistically significantly associated with haematuria across COL4A3 and COL4A4, comprising 105 nonsynonymous and 26 synonymous variants. Synonymous variants were not considered further given the lack of confidence in ascribing a plausible functional mechanism. Among the 105 nonsynonymous variants, glycine substitutions within the collagenous domain were the most frequent class (33; 34.7%), with 16 specifically substituting arginine in the Gly-X-Y motif. NC1 domain missense variants comprised 24 (25.2%) and truncating variants comprised 10 (10.5%). Domain position and effect sizes for all protein-altering variants meeting both statistical significance and pathogenicity criteria are shown in Figure 1.

**Figure 1.**
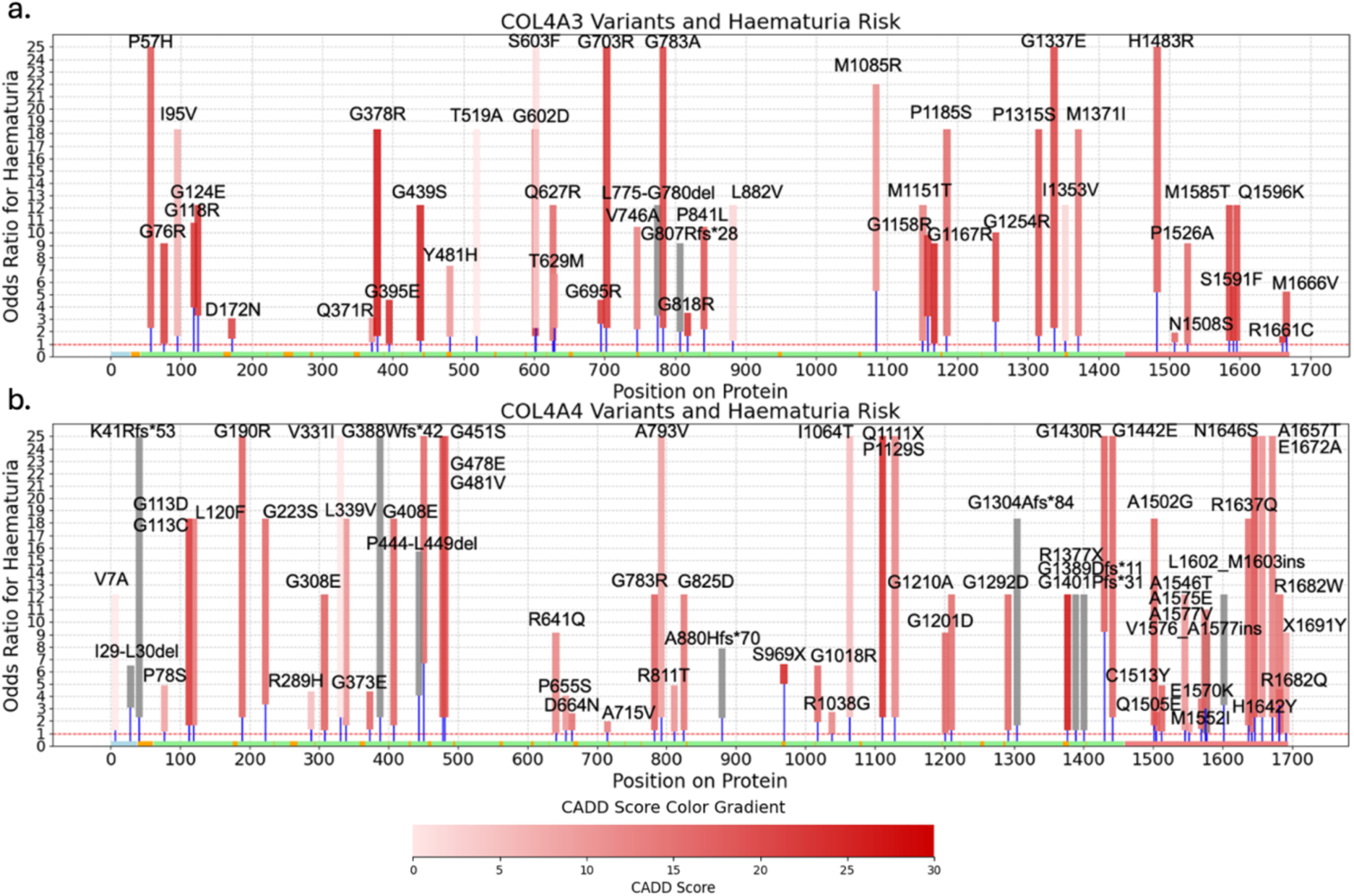
COL4A3/A4 Variants with 95% Cls Entirely above 1 for Glomerular Haematuria. Protein-altering variants with 95% confidence intervals entirely above 1 are mapped to their respective protein domains. Rectangles represent individual variants; height reflects the OR range from the lower bound of the 95% CI to the point estimate. For visualisation purposes, ORs are capped at 25, variants exceeding this threshold are displayed at the cap. Variants are colour-coded by CADD score (pale pink to dark red gradient indicating increasing predicted pathogenicity; grey indicates CADD not available). Protein domains are indicated below: N-terminus (blue), collagenous domain (green), NC1 domain (red), and non-collagenous interruptions (orange). The dashed red line at OR=1 denotes the threshold for statistical evidence of association. Variants with 95% CIs crossing 1 (n=1,872), synonymous variants and 2 with CIs below 1 are not shown.

### Per-Variant Association with Glomerular Proteinuria

For proteinuria a total of 16,074 cases (41.8% male) and 222,674 controls (46.1% male) were included in the analysis. Significant associations were observed for 87 COL4A3/A4 coding variants, of which 67 were nonsynonymous and 20 synonymous. Focusing on nonsynonymous variants, glycine substitutions predominated (22; 32.8%) with 14 substituting for arginine. NC1 missense variants represented 14 (20.9%) and truncating 7 (10.4%) of the total. Domain positions and effect sizes are shown in Figure 2.

**Figure 2.**
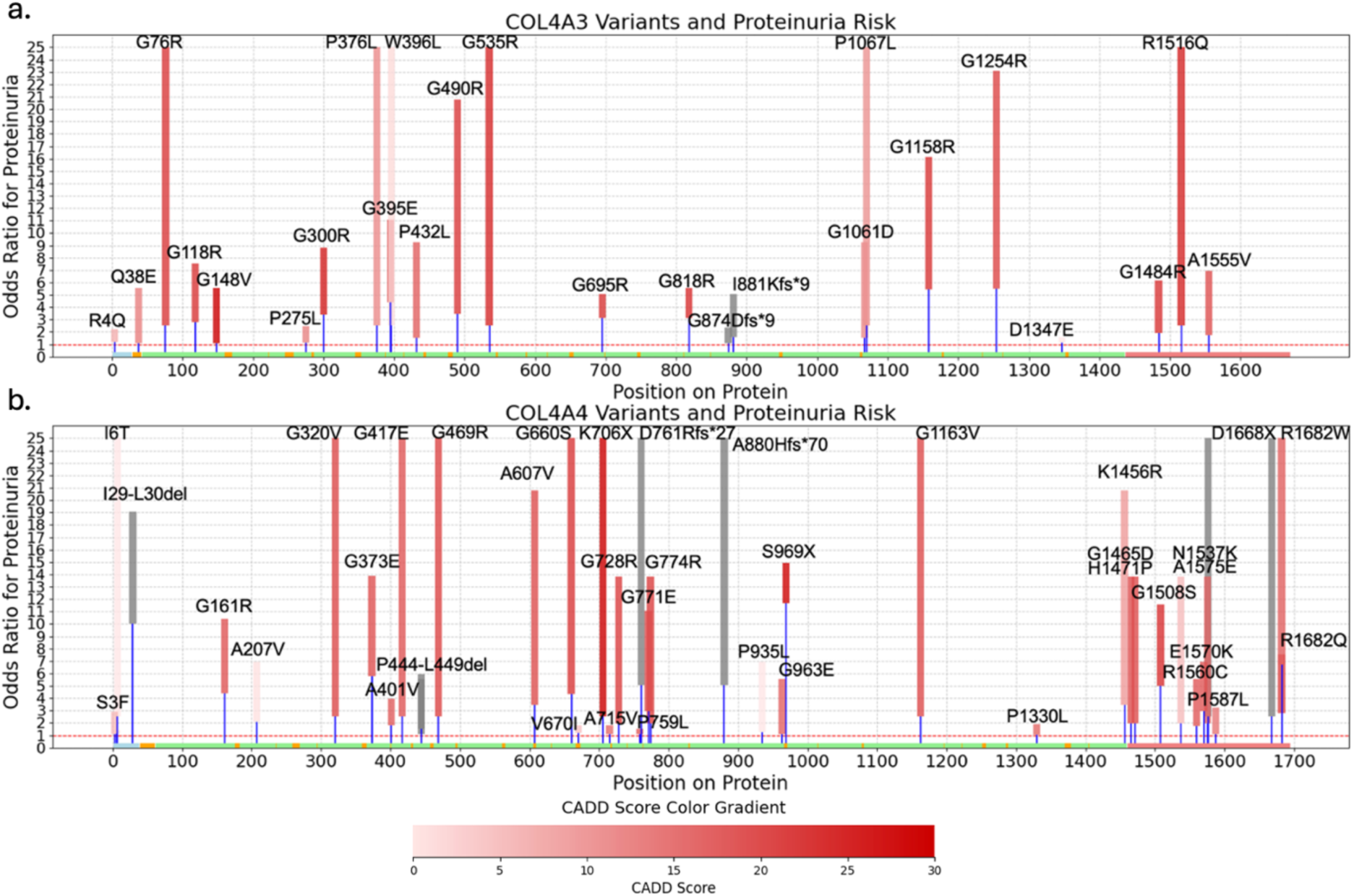
COL4A3/A4 Variants with 95% CIs Entirely above 1 for Glomerular Proteinuria. As in figure 1, for proteinuria (uACR > 3 mg/mmol). Variants with 95% crossing 1 (n=1,543), synonymous variants and 1 with CI below 1 are not shown.

### Per-Variant Association with Severe CKD

In the severe CKD analysis, 4,335 cases (61.7%) male and 293,636 controls (42.3% male) were included. Total of 62 coding variants showed statistical evidence of association, comprising 50 nonsynonymous and 12 synonymous variants. Among the nonsynonymous variants, glycine substitutions were most common (13; 26%) followed by NC1 missense (8; 16%) and truncating variants (6; 12%) (Figure 3).

**Figure 3.**
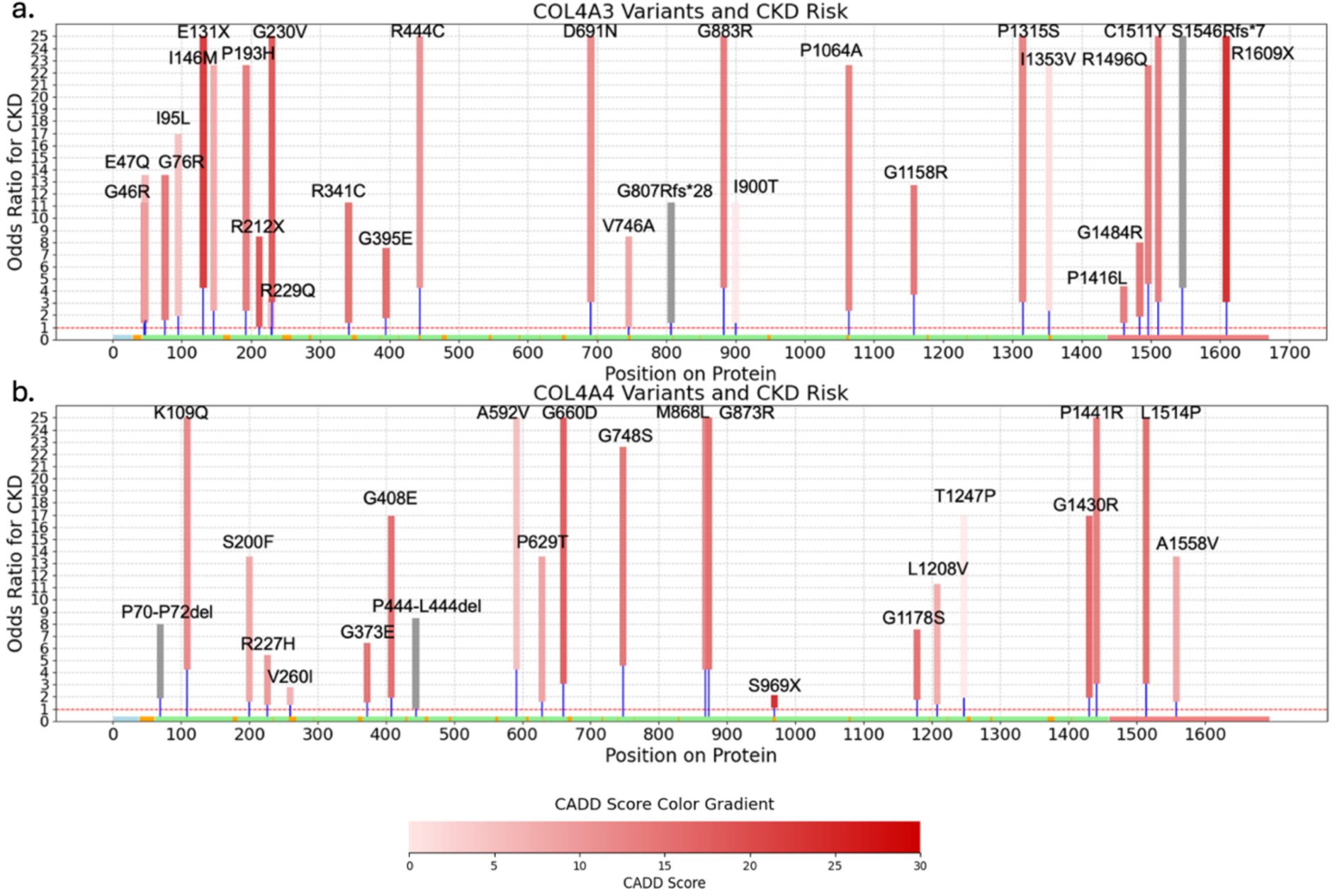
COL4A3/A4 Variants with 95% CIs Entirely above 1 for severe CKD. As in Figure 1, for severe CKD (stage 4, stage 5 and end stage renal disease). Variants with 95% crossing 1 (n=1,754) and synonymous variants are not shown.

### Comparing Carrier Case Rates between *COL4A3* and *COL4A4*

To assess carrier burden across variant classes, we examined the distribution of *COL4A3* and *COL4A4* carriers and cases for each phenotype (Table 2). COL4A4 truncating and NC1 missense variants showed consistently higher case rates than their COL4A3 counterparts across haematuria and proteinuria, with the difference most pronounced for proteinuria, where COL4A4 truncating and NC1 missense carriers had case rates of 52.7% and 37.2% respectively, compared to 17.1% and 12.1% in COL4A3. Glycine substitutions carriers showed more comparable case rates between genes for haematuria, though *COL4A4* showed higher case rates for proteinuria (48.6% vs 22.7%). For CKD, carrier numbers in most variant classes were insufficient to draw firm conclusions.

**Table 1.** Distribution of associated COL4A3 and COL4A4 variants by variant type and phenotype. Numbers indicate the count of distinct deleterious variants significantly associated with each renal phenotype, stratified by gene and variant type. Percentages represent the proportion of associated variants for that phenotype contributed by each variant class.

| Gene | Variant Type | Glomerular Haematuria | Glomerular Proteinuria | Severe CKD |
| --- | --- | --- | --- | --- |
| **COL4A3** | Total Deleterious | n = 43 | n = 25 | n = 28 |
|  | Gly-Subs | 15 (34.9%) | 11 (44%) | 6 (21.4%) |
|  | Truncating | 1 (2.3%) | 2 (8%) | 5 (17.8%) |
|  | NC1 missense | 8 (18.6%) | 3 (12%) | 6 (21.4%) |
| **COL4A4** | Total Deleterious | n = 62 | n = 42 | n = 22 |
|  | Gly-Subs | 18 (29.1%) | 11 (26.2%) | 7 (31.8%) |
|  | Truncating | 9 (14.5%) | 5 (11.9%) | 1 (4.5%) |
|  | NC1 missense | 16 (25.8%) | 11 (26.2%) | 2 (9.1%) |

| **COL4A3/A4** | Total Deleterious | n = 95 | n = 67 | n = 50 |
| --- | --- | --- | --- | --- |
|  | Gly-Subs | 33 (34.7%) | 22 (32.8%) | 13 (26%) |
|  | Truncating | 10 (10.5%) | 7 (10.4%) | 6 (12%) |
|  | NC1 missense | 24 (25.2%) | 14 (20.9%) | 8 (16%) |

**Table 2.** Carrier counts for COL4A3 and COL4A4 variants by variant class and phenotype in the UK Biobank. Numbers indicate the total count of carriers for each variant class, with cases in parentheses. Carrier counts of variants outside of the three predefined variant classes are not shown.

| **Gene** | **Phenotype** | **All carriers**<br>**(cases;%)** | **Glycine substitutions**<br>**(cases;%)** | **Truncating**<br>**(cases;%)** | **NC1 missense**<br>**(cases;%)** |
| --- | --- | --- | --- | --- | --- |
| **COL4A3** | Haematuria | 1512 (133;8.8%) | 317 (46;14.5%) | 10 (2;20%) | 945 (50;5.3%) |
|  | Proteinuria | 975 (172;17.6%) | 388 (88;22.7%) | 70 (12;17.1%) | 372 (45;12.1%) |
|  | CKD | 282 (37;13.1%) | 57 (9;15.8%) | 23 (5;21.7%) | 79 (8;10.1%) |
| **COL4A4** | Haematuria | 2083 (186;8.9%) | 104 (26;25%) | 430 (70;16.3%) | 573 (46;8%) |
|  | Proteinuria | 1465 (354;24.2%) | 107 (52;48.6%) | 275 (145;52.7%) | 121 (45;37.2%) |
|  | CKD | 603 (38;6.3%) | 66 (10;15.2%) | 291 (9;3.1%) | 9 (2;22.2%) |

**Table 3.**
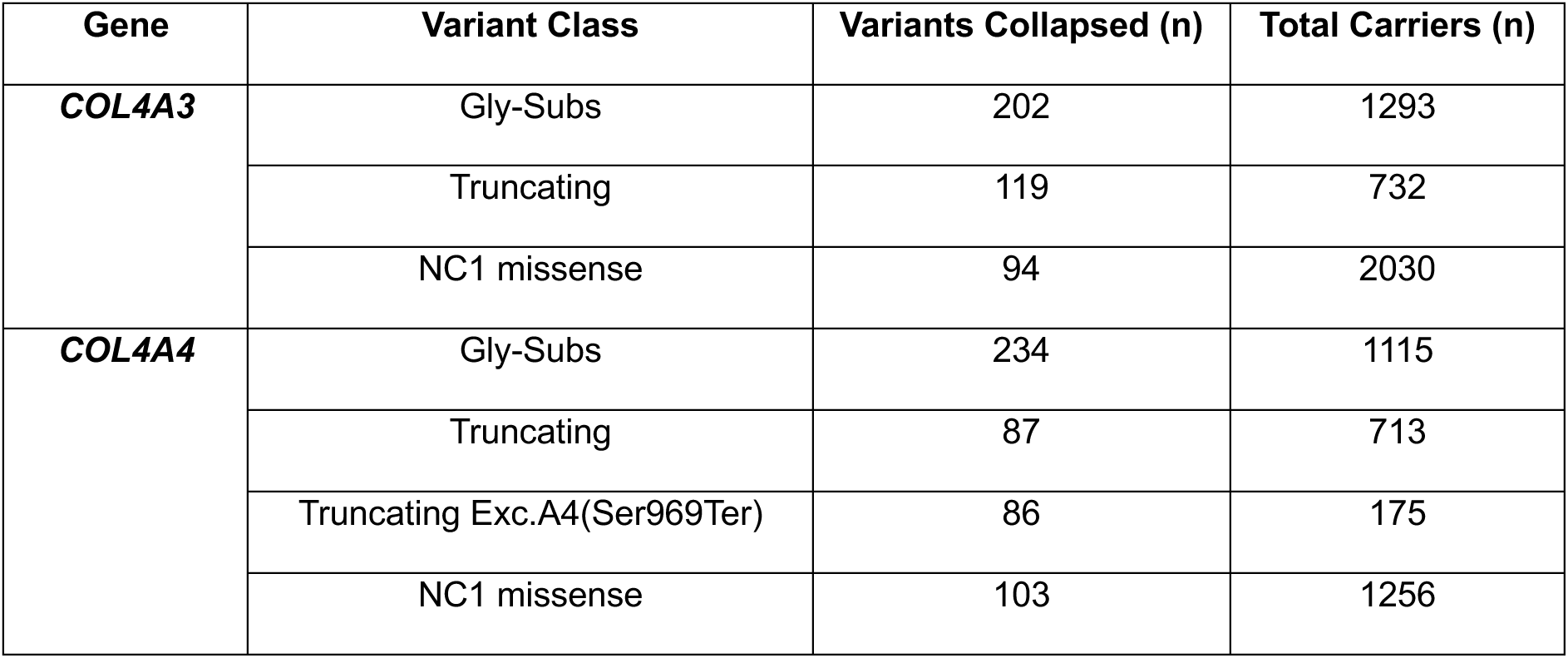
Summary of collapsed variant counts and carrier frequencies by gene and variant class in UKB. For each gene (COL4A3, COL4A4) and variant class (glycine substitutions, truncating variants, NC1 missense variants), the total number of rare variants collapsed and the total number of carriers in the UK Biobank White European subset are shown.

| Gene | Variant Class | Variants Collapsed (n) | Total Carriers (n) |
| --- | --- | --- | --- |
| COL4A3 | Gly-Subs | 202 | 1293 |
|  | Truncating | 119 | 732 |
|  | NC1 missense | 94 | 2030 |
| COL4A4 | Gly-Subs | 234 | 1115 |
|  | Truncating | 87 | 713 |
|  | Truncating Exc.A4(Ser969Ter) | 86 | 175 |
|  | NC1 missense | 103 | 1256 |

**Table 4.** Summary of collapsed variant counts and carrier frequencies by gene and variant class in All of Us. For each gene (COL4A3, COL4A4) and variant class (glycine substitutions, truncating variants, NC1 missense variants), the total number of rare variants collapsed and the total number of carriers in the All of Us White European subset are shown.

| Gene | Variant Class | Variants Collapsed (n) | Total Carriers (n) |
| --- | --- | --- | --- |
| **COL4A3** | Gly-Subs | 155 | 515 |
|  | Truncating | 82 | 136 |
|  | NC1 missense | 81 | 852 |
| **COL4A4** | Gly-Subs | 162 | 666 |
|  | Truncating | 61 | 116 |
|  | NC1 missense | 98 | 667 |

### Collapsing PheWAS: Glycine Substitutions

To assess the aggregate phenotypic burden of rare glycine substitutions in the collagenous domain, we performed a PheWAS collapsing rare (MAF < 1%) *COL4A3* and *COL4A4* gly-sub variants separately, testing each gene against 1,363 ICD-10 phenotypes, including the Proteinuria uACR > 3 mg/mmol and severe CKD previously defined phenotypes, in the UK Biobank (Figure 4). A total of 202 *COL4A3* gly-sub variants (carrier count: n = 1,293) and 234 *COL4A4* gly-sub variants (carrier count: n = 1,115) were collapsed into binary variables. For *COL4A3*, the strongest associations were observed with proteinuria (uACR > 3 mg/mmol: β = 0.04, p = 2.71 × 10^−57^) and unspecified haematuria (R31; β = 0.114, p = 4.97 × 10^−18^). Additionally significant associations included isolated proteinuria (R80; β = 0.114, p = 4.97 × 10^−7^) and unspecified kidney disorders (N28; β = 0.033, p = 1.78 × 10^−6^). For *COL4A4*, gly-sub carriers also showed highly significant associations with proteinuria (uACR > 3 mg/mmol: β = 0.054, p = 3.08 × 10^−32^) and unspecified haematuria (R31; β = 0.039, p = 1.19 × 10^−15^). In addition, recurrent and persistent haematuria (N02; β = 0.076, p = 3.70 × 10^−6^) and unspecified kidney disorders (N28; β = 0.03, p = 2.58 × 10^−5^) reached significance. No other phenotype classes showed consistent significant associations after correction for multiple testing.

**Figure 4.**
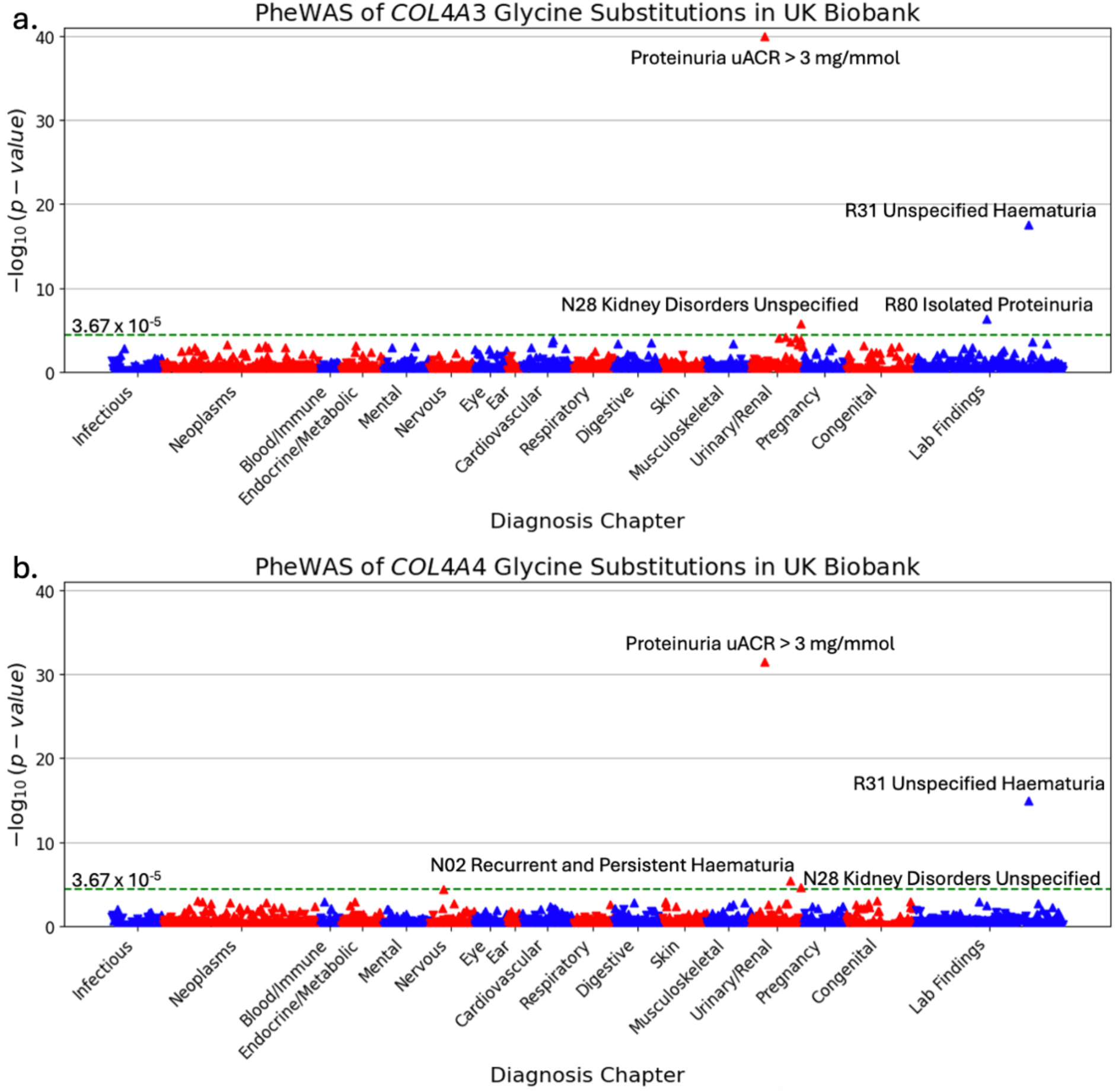
Manhattan plots of diagnoses for collapsing tests of COL4A3/A4 glycine substitutions in UK Biobank. Manhattan plot showing the association of diagnoses with COL4A3 glycine substitutions. b. Manhattan plot diagnoses associated with glycine substitutions in COL4A4. For visualisation purposes, -log10(p-values) are capped at 40, associations exceeding this threshold are displayed at the cap. Diagnoses are grouped by phenotypic categories on the x-axis. Upward-pointing triangles represent positive associations (OR > 1), while downward-pointing triangles represent negative associations (OR < 1). Red dashed line indicates the Bonferroni-corrected significance threshold.

### Collapsing PheWAS: Truncating Variants

We next performed a PheWAS collapsing rare, high-confidence loss-of-function (truncating) variants in *COL4A3* and *COL4A4* separately, aggregating 119 variants in *COL4A3* (n = 732 carriers) and 87 in *COL4A4* (n = 713 carriers) into binary burden variables (Figure 5). *COL4A3* truncating variants showed no significant associations after Bonferroni correction; the strongest signal was observed for proteinuria (uACR > 3 mg/mmol; β = 0.031, p = 9.51 × 10^−5^), which fell below the significance threshold (p < 3.67 × 10^−5^). In contrast, *COL4A4* truncating variants were highly significantly associated with a broad range of kidney phenotypes. The strongest associations were observed with proteinuria (uACR > 3 mg/mmol; β = 0.107, p = 1.39 × 10^−119^) and unspecified haematuria (R31; β = 0.083, p = 8.26 × 10^−55^). Additional significant associations included recurrent and persistent haematuria (N02; β = 0.112, p = 1.75 × 10^−12^), unspecified kidney disorders (N28; β = 0.045, p = 5.10 × 10^−8^), chronic renal failure (N18; β = 0.022, p = 9.51 × 10^−6^) and gout (M10; β = 0.018, p = 4.83 × 10^−6^).

**Figure 5.**
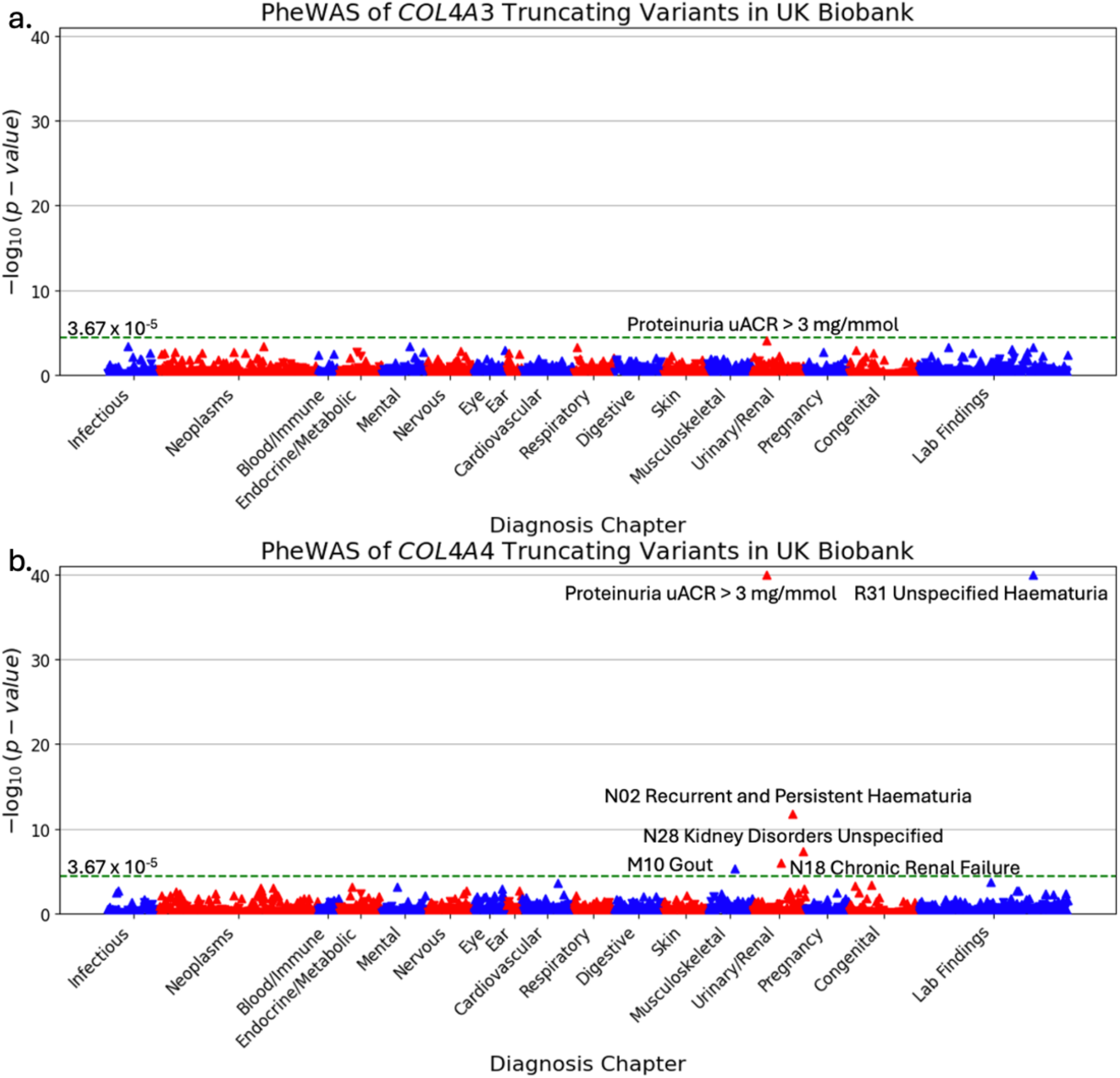
Manhattan plots of diagnoses for collapsing tests of COL4A3/A4 truncating variants in UK Biobank. a. COL4A3 truncating variants. b. COL4A4 truncating variants. See Figure 4 legend for detailed description of plot elements.

### Sensitivity Analysis: Effect of p.Ser969Ter on *COL4A4* Truncating Signal

To determine whether the strong phenotypic signal observed for *COL4A4* truncating variants was disproportionately driven by a single high-frequency founder variant endemic in the English population, we performed a sensitivity analysis repeating the collapsing PheWAS for *COL4A4* truncating variants after excluding the p.Ser969Ter variant (rs144624277, NM_000092.5:c.2906C>A), which is known to be enriched in individuals of British ancestry (MAF = 0.13%). This reduced the collapsed variant set from 87 to 86 truncating variants and the total carrier count from 713 to 175 carriers. Despite the substantial reduction in carrier number, the association with proteinuria (uACR > 3 mg/mmol; β = 0.098, p = 9.51 × 10^−22^) and unspecified haematuria (R31; β = 0.087, p = 9.12 × 10^−17^) remained highly significant and of similar magnitude. However, all other previously associated phenotypes, including recurrent haematuria (N02), unspecified kidney disorders (N28), chronic renal failure (N18) and gout (M10), no longer reached Bonferroni-corrected significance (Figure 6). These findings indicate that while p.Ser969Ter contributes to the breadth of the *COL4A4* truncating signal, the core associations with glomerular haematuria and proteinuria are robust and not purely attributable to this single variant.

**Figure 6.**
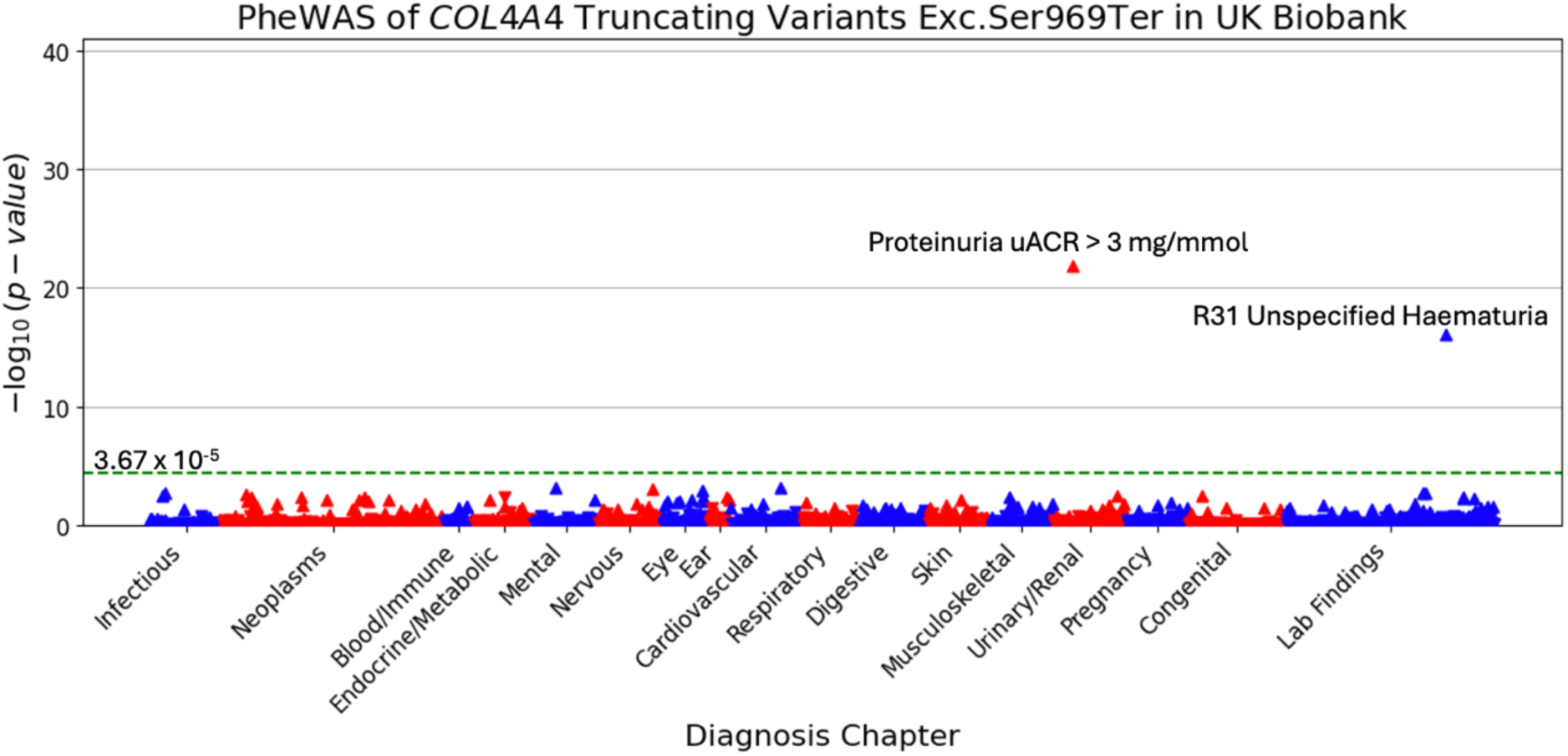
Manhattan plots of diagnoses for collapsing tests of COL4A4 truncating variants excluding p.Ser969Ter. Manhattan plot showing association of diagnoses with COL4A4 truncating variants after removal of the British founder variant p.Ser969Ter. See Figure 4 legend for detailed description of plot elements.

### Collapsing PheWAS: NC1 Missense Variants

We then conducted a PheWAS that collapsed rare, pathogenic NC1 domain missense variants (CADD > 20) into binary burden variables for *COL4A3* (94 variants; 2,030 carriers) and *COL4A4* (103 variants; 1,256 carriers) individually (Figure 7). Replicating the pattern observed for truncating variants, *COL4A3* NC1 missense variants showed no significant associations; the strongest signal was for unspecified haematuria (R31; β = 0.0095, p = 4.81 × 10^−4^). In contrast, *COL4A4* NC1 missense variants were significantly associated with both proteinuria (uACR > 3 mg/mmol; β = 0.042, p = 5.51 × 10^−22^) and unspecified haematuria (R31; β = 0.025, p = 9.46 × 10^−7^).

**Figure 7.**
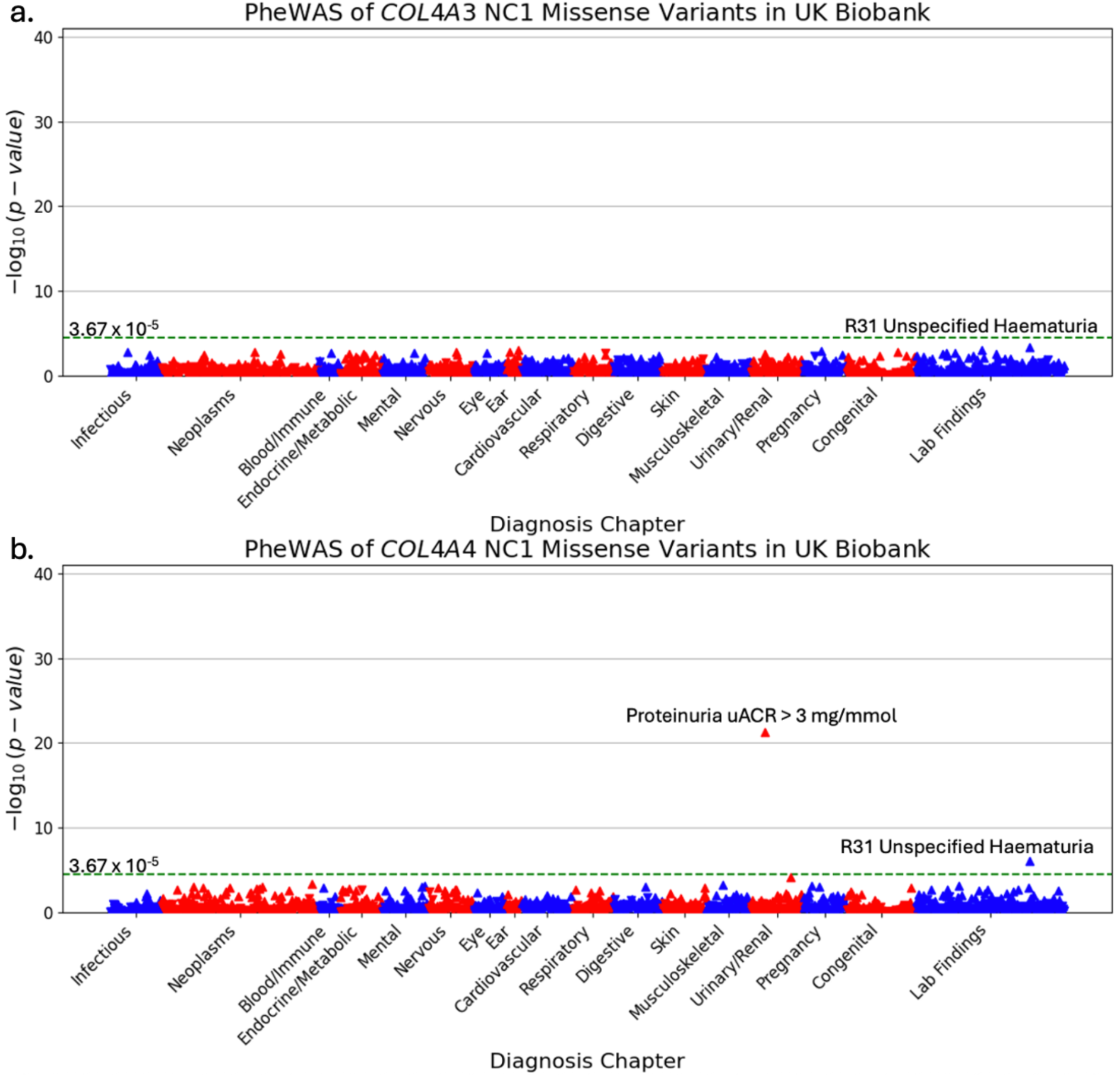
Manhattan plots of diagnoses for collapsing tests of COL4A3/A4 NC1 domain missense variants in UK Biobank. a. COL4A3 NC1 missense variants. b. COL4A4 NC1 missense variants. See Figure 4 legend for detailed description of plot elements.

### Comparison of Genetic Effect Sizes between *COL4A3* and *COL4A4*

To formally test whether the phenotype impact of rare variants differs between *COL4A3* and *COL4A4*, we compared the effect size estimates (BETAs) from the SAIGE PheWAS models for haematuria and proteinuria using two-tailed Z-tests, performed separately for each variant class (Figure 8). Glycine substitutions showed equivalent effect sizes between the two genes for both haematuria (Δβ = −0.0022, Z = 0.323, p = 0.746) and proteinuria (Δβ = - 0.0054, Z = −0.855, p = 0.392). In striking contrast, truncating variants in *COL4A4* exhibited significantly larger effect sizes than their *COL4A3* counterparts for both haematuria (Δβ = - 0.0822, Z = −8.49, p = 5.12 × 10^−18^) and proteinuria (Δβ = −0.0804, Z = −10.17, p = 6.92 × 10^−25^). This gene-specific difference persisted after exclusion of the British founder variant p.Ser969Ter from the *COL4A4* truncating model, with *COL4A4* variants remaining significantly more impactful than *COL4A3* truncating variants for haematuria (Δβ = −0.0761, Z = −5.73, p = 1.01 × 10^−8^) and proteinuria (Δβ = −0.0404, Z = −6.87, p = 6.63 × 10^−12^). This divergence was also observed despite *COL4A3* NC1 missense variants having substantially more carriers than *COL4A4* (n = 2,030 vs. n = 1,256). Despite similar numbers of collapsed variants and carrier counts between the two genes across all three variant classes (Table 2) and comparable CADD scores for *COL4A3* and *COL4A4* NC1 missense variants (mean CADD: 24.6 vs 23.9, respectively) (Supplementary Figure S4), these findings demonstrated a consistent gene-specific effect: truncating and NC1 domain variants confer significantly greater risk when located in *COL4A4*, while glycine substitutions confer equivalent risk irrespective of the affected gene.

**Figure 8.**
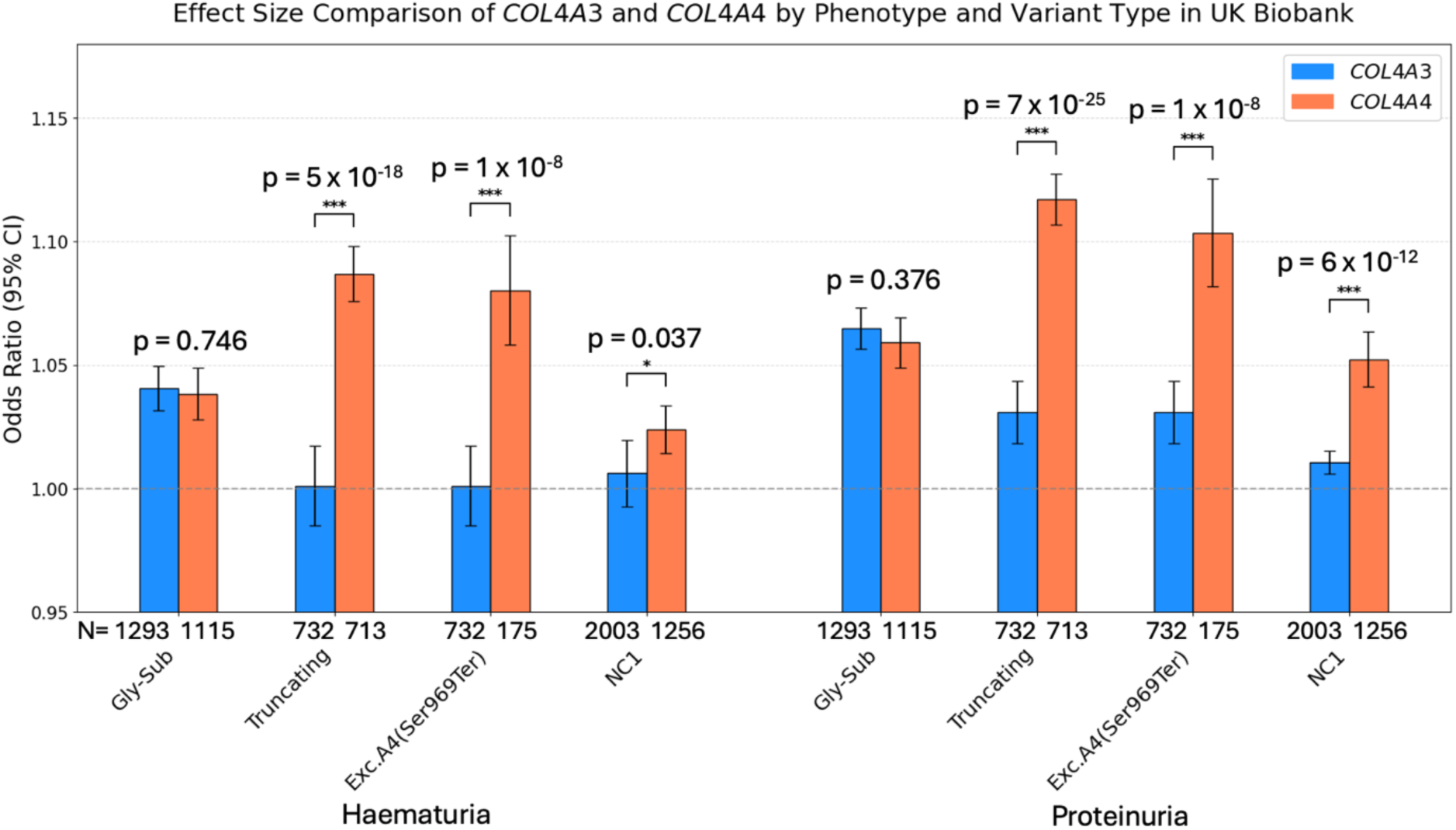
Comparison of ORs for haematuria and proteinuria between COL4A3 and COL4A4 by variant type in UK Biobank. Comparing ORs for haematuria and proteinuria between COL4A3 and COL4A4, stratified by variant class (Gly-sub, Truncating, NC1). Error bars show 95% CIs, Carrier count (N) is shown for each group. P-values calculated via two-tailed Z-tests.

### Replication of PheWAS in All of Us Cohort

We replicated the collapsing PheWAS analyses in the All of Us Research Program cohort (v8), restricted to individuals of European ancestry aged ≥40 years (n ≈ 160,000), using identical variant classification and statistical methods (Bonferroni threshold p < 3.8 × 10^−5^). The six collapsing models were tested against 1,322 ICD-10 phenotypes (Supplementary Figures S6-S8). Glycine substitutions in both genes replicated the associations with haematuria observed in UK Biobank. *COL4A3* gly-sub carriers showed significant associations with unspecified (R31; β = 0.025, p= 2.78 × 10^−6^) and recurrent/persistent haematuria (N02; β = 0.107, p= 2.94 × 10^−5^). *COL4A4* gly-sub carriers were also significantly associated with both R31 (β = 0.029, p= 4.34 × 10^−14^) and N02 (β = 0.112, p= 1.54 × 10^−5^). Truncating variants recapitulated the gene-specific pattern observed in UKB with *COL4A3* truncating variants showed no significant associations (strongest signal: R31, p= 9.10 × 10^−4^) whereas *COL4A4* truncating variants were significantly associated with R31 (β = 0.064, p= 2.24 × 10^−10^) and unspecified kidney disorders (N28; β = 0.052, p= 1.11 × 10^−5^). NC1 missense variants similarly showed no significant associations for *COL4A3* (strongest signal: R31, p= 2.91 × 10^−4^), while *COL4A4* NC1 variants were significantly associated with R31 (β = 0.024 p= 5.86 × 10^−7^).

Notably, while proteinuria was among the top associations for most models in All of Us, ranking second or third after haematuria phenotypes, it did not reach Bonferroni significance in any analysis. This may reflect differences in laboratory data availability, measurement protocols, or population structure between cohorts and is discussed further in Supplementary Results (Supplementary Note S6). Nevertheless, effect size estimates for proteinuria were extracted from all models to enable formal comparison between genes and across cohorts.

When comparing effect sizes between *COL4A3* and *COL4A4* in the All of Us replication cohort using two-tailed Z-tests, for haematuria the pattern observed in UKB was largely recapitulated (Figure 9). Glycine substitutions showed equivalent effects between genes (Δβ = −0.0046, Ζ = −0.65, p = 0.514), while truncating *COL4A4* conferred significantly greater risk for haematuria than those in *COL4A3* (Δβ = −0.0630, Ζ = −2.40, p = 0.0163). Similarly, NC1 missense variants in *COL4A4* demonstrated a significantly larger effect than their *COL4A3* counterparts (Δβ = −0.0180, Ζ = −2.85, p = 0.0044). For proteinuria, glycine substitutions again showed no difference between genes (Δβ = 0.0046, Ζ = 0.25, p = 0.806). Truncating variants in *COL4A4* remained significantly more impactful than those in *COL4A3* (Δβ = - 0.0481, Ζ = −1.96, p = 0.0499), consistent with the UK Biobank finding despite the substantially lower frequency for the British founder variant p.Ser969Ter in All of Us (MAF = 0.0094% vs 0.13% in UKB; AC = 77). The gene-specific difference for NC1 missense variants did not replicate for proteinuria in All of Us (Δβ = −0.0173, Ζ = −1.21, p = 0.225). These findings broadly confirm the gene-specific effect observed in UK Biobank, particularly for haematuria, with *COL4A4* truncating and NC1 variants conferring greater risk than their *COL4A3* equivalents.

**Figure 9.**
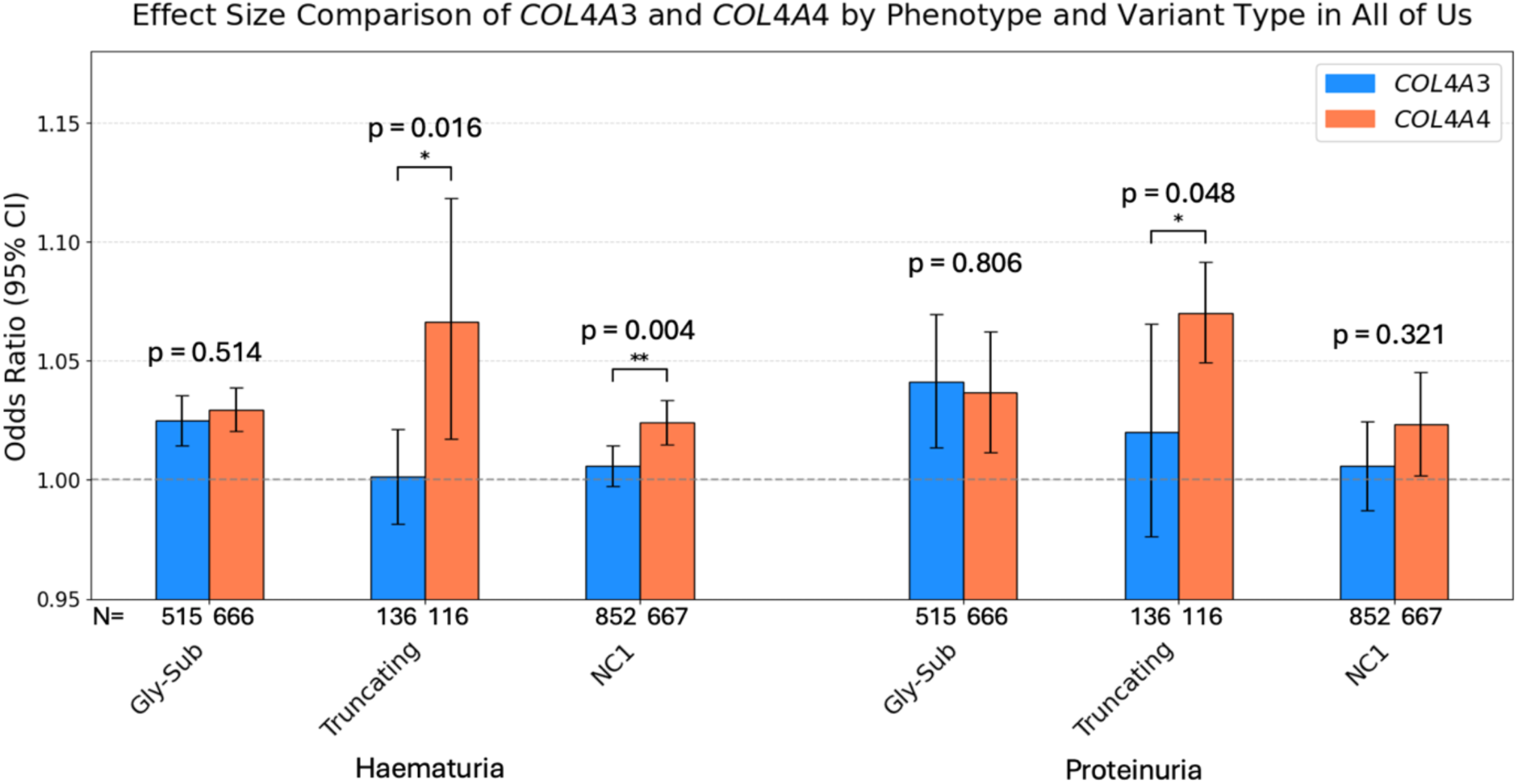
Comparison of ORs for haematuria and proteinuria between COL4A3 and COL4A4 by variant type in All of Us. Comparing ORs for haematuria and proteinuria between COL4A3 and COL4A4, stratified by variant class (Gly-sub, Truncating, NC1). Error bars show 95% CIs, Carrier count (N) is shown for each group. P-values calculated via two-tailed Z-tests.

### Genome-Wide Association Study for Haematuria

To identify genetic loci associated with glomerular haematuria, we performed a GWAS in the UK Biobank with 8,224 cases and 41,120 matched controls. The *COL4A3/A4* locus on chromosome 2 was the strongest association signal genome-wide, with the lead variant being the *COL4A4* p.Ser969Ter founder variant (rs144624277; β = 3.34, p = 1.8 × 10^−23^) (Figure 10a). These findings were broadly consistent with a previous haematuria GWAS conducted in this dataset^17^. In addition to this primary signal, we observed significant association at the *THSD4* locus on chromosome 15, where four intronic SNPs approached genome-wide significance, replicating a reported association with haematuria in the Million Veteran Program^18^. The *HLA-B* region on chromosome 6 also reached genome-wide significance, consistent with previous GWAS of haematuria^17,19^. Colocalization analysis with a GWAS of IgA nephropathy^20^ identified a shared signal at rs521977 (PP.H4 = 0.99; Supplementary Figure S12), a variant in linkage disequilibrium with HLA-B*08:01, which was previously shown to fully explain the *HLA-B* haematuria signal by conditional analysis^17^.

**Figure 10.**
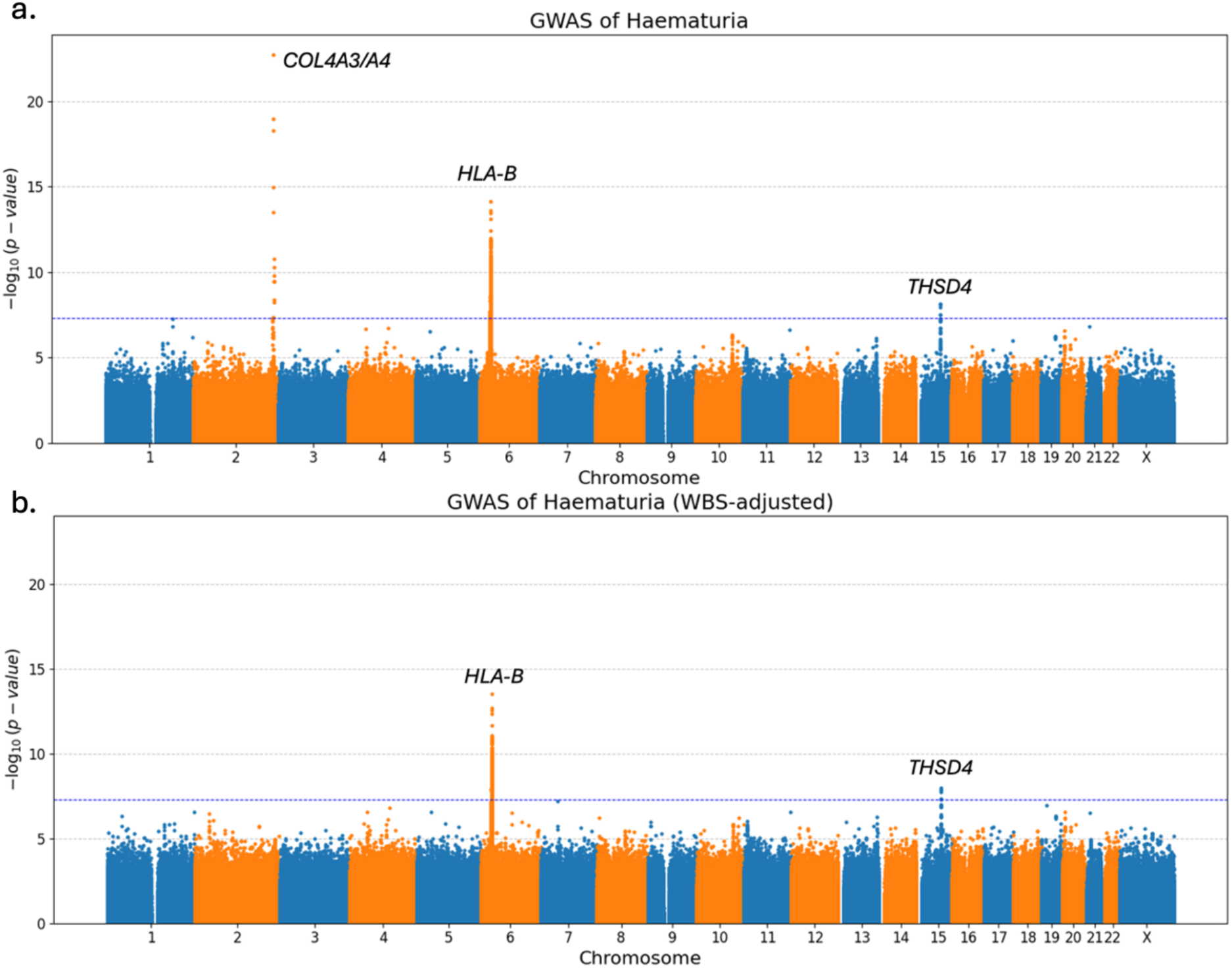
Genome-wide association studies for haematuria. a. Manhattan plot showing association strength (-log10 P-value) across the genome for haematuria (λ = 1.029). b. Manhattan plot after including WBS as a covariate (λ = 1.021). Red line indicates genome-wide significance threshold (p < 5 × 10^−8^).

To determine whether the *COL4A3/A4* signal was driven by the aggregate effect of rare variants and to identify potential trans-acting modifiers independent of the primary locus, we performed a second GWAS including a weighted *COL4A3/A4* burden score (WBS) as a continuous covariate (Figure 10b). The WBS was calculated for each individual as the sum per-variant ORs for haematuria multiplied by genotype dosage, restricted to variants showing statistical evidence of association with haematuria in the per-variant analysis. After adjusting for WBS, the association at the *COL4A3/A4* locus was completely abolished, indicating that the signal can be attributed to the aggregate effect of rare pathogenic variants in these genes. Both *HLA-B* and *THSD4* remained genome wide significant after adjustment. No new loci reached genome-wide significance in the adjusted model, indicating absence of strong trans-acting modifiers of haematuria risk in this population.

### GWAS for Glomerular Haematuria Restricted to *COL4A3/A4* Heterozygotes

We performed a secondary GWAS restricted to heterozygotes for any qualifying *COL4A3* or *COL4A4* variant (glycine substitutions, truncating and NC1 domain missense variants with CADD > 20) to investigate genetic modifiers operating specifically in these individuals (1,627 cases, 8,755 controls). The *COL4A3/A4* locus remained the dominant signal, with the lead variant being the *COL4A4* p.Ser969Ter founder variant (rs144624277; β = 4.15, p = 3.3 × 10^−21^) (Figure 11a). After adjusting for the WBS, the *COL4A3/A4* signal was abolished (Figure 11b).

**Figure 11.**
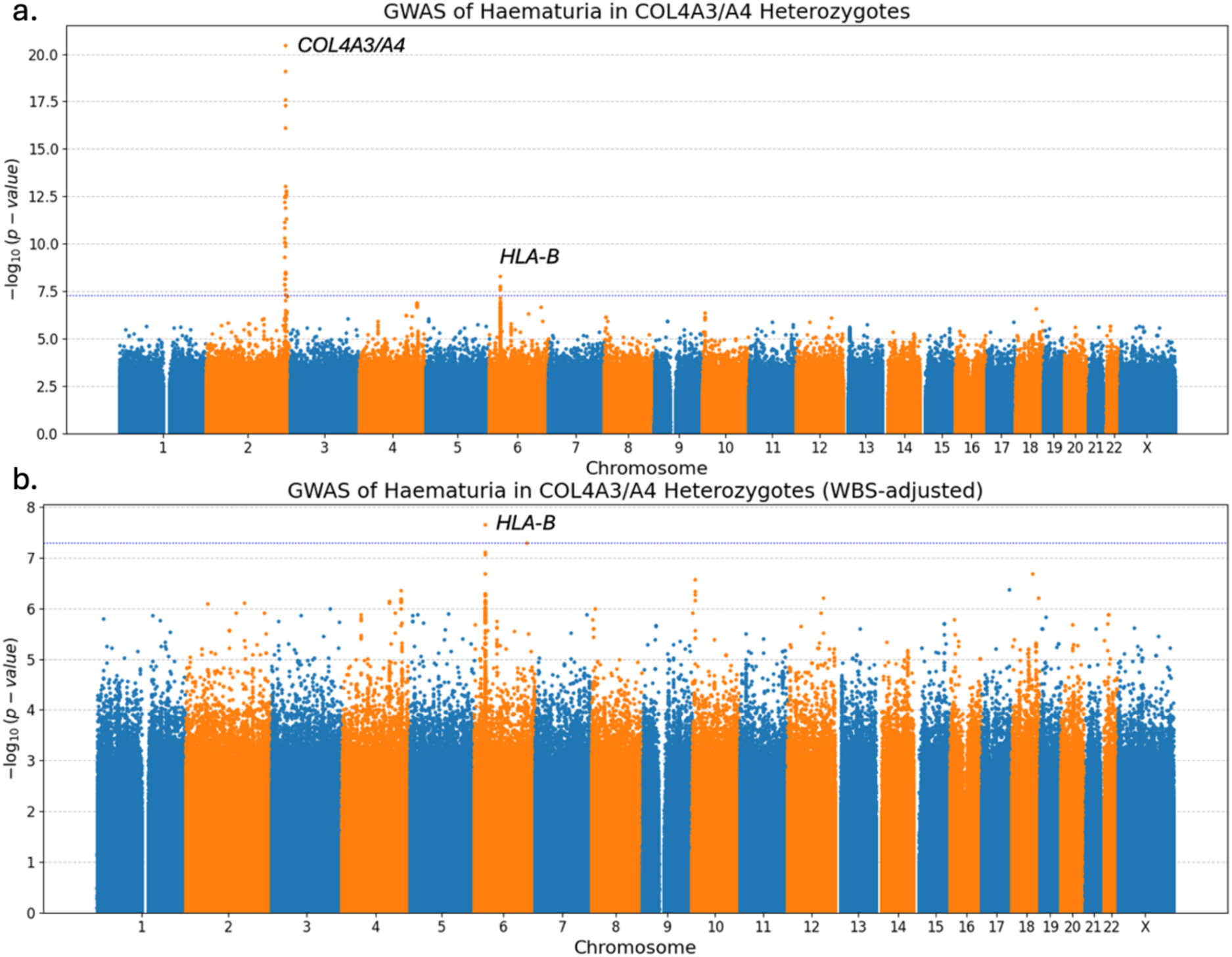
Genome-wide association studies for glomerular haematuria restricted to COL4A3/A4 heterozygotes. a. Manhattan plot showing association strength (-log10 P-value) across the genome for glomerular haematuria in individuals carrying a qualifying COL4A3 or COL4A4 variant (λ = 1.031). b. Manhattan plot after including WBS as a covariate (λ = 1.021). Red line indicates genome-wide significance threshold (p < 5 × 10^−8^).

In the unadjusted analysis, the HLA-B region on chromosome 6 was the only other locus to reach genome-wide significance and remained significant after the adjustment for WBS. No other loci reached genome-wide significance in either analysis.

## Discussion

In this study, we leveraged large-scale population biobanks to quantify the penetrance and disease burden of rare *COL4A3* and *COL4A4* variants in an unselected population, avoiding the ascertainment bias of clinically recruited cohorts.

Our per-variant analysis identified 105 and 70 rare variants significantly associated with haematuria and proteinuria, respectively, generating a quantitative allelic series that can inform clinical variant classification and enable adjustment for primary mutation severity in future studies. Glycine substitutions were the most frequent variant class across all renal phenotypes, consistent with their established role in disrupting collagen triple helix formation^21,22^.

Strikingly, we uncovered a strong gene-specific effect: truncating and NC1 domain missense variants were primarily pathogenic in *COL4A4*, while *COL4A3* exhibited greater tolerance to these variant classes, evidenced by the greater number of variant carriers and reduced evidence of association with phenotypes. This divergence persisted despite similar numbers of collapsed variants, comparable carrier counts, and equivalent CADD scores between the two genes, and remained after excluding the British founder variant p.Ser969Ter. In contrast, glycine substitutions showed equivalent effect sizes in *COL4A3* and *COL4A4*, indicating that the gene-specific effect is confined to loss-of-function and NC1-domain variants. These findings suggest that trimer formation or function may be less dependent on collagen α3(IV) than α4(IV) protein structure or abundance, mitigating the impact of truncating and NC1 variants in *COL4A3*, challenging an assumption of functional equivalence between the proteins encoded by these two genes. These findings have implications beyond Alport syndrome because they demonstrate that paralogous genes encoding subunits of the same heteromeric complex can differ substantially in their tolerance to loss-of-function variants, and that variant class alone is insufficient to predict pathogenicity without considering which paralogue is affected. This principle may apply to other heterotrimer-encoding gene families and argues for gene-aware rather than variant-class-aware frameworks in clinical variant interpretation.

Finally, we show that the *COL4A3/A4* locus is the dominant genetic signal for glomerular haematuria, and the signal can be attributed to the aggregate effect of rare variants. After adjusting for a weighted burden score, the locus signal was abolished and no significant trans-acting modifiers emerged, suggesting that the genetic architecture of haematuria is largely explained by the direct effects of rare variants in these genes, without evidence of independent trans-acting modifiers. This is further supported by the secondary GWAS restricted to *COL4A3/A4* heterozygotes where the *COL4A3/A4* locus remained the dominant signal. After adjustment for the WBS, this signal was abolished, consistent with the primary GWAS findings. The per-variant ORs used to construct the WBS were estimated without conditioning on local haplotype structure since most variants are too rare to allow this. For higher-frequency rare variants such as the British founder variant *COL4A4:* p.Ser969Ter, it is possible that some of the attributed effect reflects linkage to common variants on the ancestral haplotype rather than the coding variant itself. The abolition of the *COL4A3/A4* GWAS signal after WBS adjustment should therefore be interpreted as showing that the signal is accounted for by the aggregated per-variant effects, not necessarily that common variant effects at this locus are absent.

The persistence of the *HLA-B* signal in this GWAS raises the intriguing possibility that immunological mechanisms might contribute to risk of kidney disease among individuals harbouring a single pathogenic *COL4A3* or *COL4A4* variant. In addition, the *THSD4* and *HLA-B* loci were identified as independent loci associated with haematuria, replicating previously reported association with haematuria and remained genome-wide significant after adjustment. Colocalization analysis suggests that the *HLA-B* association reflects contributions from immune-mediated glomerular diseases including IgA nephropathy, which are strongly HLA-associated and may co-occur independently with *COL4A3/A4* carrier status. Although an immunological contribution to disease risk in *COL4A3/A4* carriers cannot be excluded, this signal should not be over-interpreted as evidence of a specific gene-immune interaction without replication in a cohort with granular glomerular phenotyping (i.e. that has undergone kidney biopsy examination).

Our analyses were restricted to individuals of European ancestry, and findings may not generalise to other populations – among which additional (as yet unascertained) genetic associations may exist. Phenotype definitions relied on ICD-10 codes and available laboratory measurements, which may underestimate true prevalence since ICD-10 codes capture haematuria that has generated a hospital episode. Asymptomatic microscopic haematuria, which is the cardinal early manifestation of *COL4A3/A4*-associated disease, is typically identified in primary care and is unlikely to generate the ICD-10 codes used here unless a referral to secondary care is made. The UK Biobank is a healthy volunteer cohort, which further biases towards under-representation of (severe) disease in variant carriers. The approach to identification of haematuria taken therefore maximises specificity at the cost of sensitivity, and carrier case rates derived from these data should be interpreted as conservative estimates of penetrance. In All of Us, proteinuria is not uniformly defined, potentially attenuating replication of genetic associations with this trait. Finally, the biochemical or structural mechanisms underlying the gene-specific effects we observed remain to be elucidated. An intriguing possibility is that the divergent genetic associations observed for *COL4A3* and *COL4A4* arise from non-equivalent roles of the encoded chains during α3α4α5(IV) heterotrimer assembly. Previous structural and biochemical studies indicate that specific NC1-domain interactions govern chain recognition and assembly of the mature collagen IV protomer, suggesting that the individual chains may contribute differently to this process^23–25^. One potential explanation is that α4(IV) plays a disproportionately important role in chain selection or protomer stabilization, rendering truncating and NC1-domain variants of *COL4A4* less well tolerated than those of *COL4A3*. However, this possibility remains speculative because the specific contribution of the α4(IV) NC1 domain to chain-selection dynamics has not yet been directly examined.

In conclusion, this population-based study provides per variant quantitative evidence that rare *COL4A3/A4* variants are a significant cause of haematuria and proteinuria. We demonstrate that truncating and NC1 domain variants in *COL4A4* have a substantially stronger pathogenic effect than their counterparts in *COL4A3*, while glycine substitutions confer equivalent risk in both genes. The *COL4A3/A4* locus is the dominant signal for glomerular haematuria, which is driven entirely by the quantified effects of rare variants. These findings refine the genetic architecture of collagen IV nephropathies and highlight the need for gene-aware approaches to clinical management. More broadly, these data demonstrate that paralogous genes encoding subunits of the same protein complex can differ substantially in their sensitivity to loss-of-function variation, and that variant class alone is insufficient to predict pathogenicity without considering gene identity — a principle with potential relevance to clinical interpretation across other heteromultimer-encoding gene families.

## Data Availability

UK Biobank data are available to approved researchers through the UK Biobank Access Management System (https://www.ukbiobank.ac.uk). All of Us Research Program data are available to registered researchers through the All of Us Researcher Workbench (https://www.researchallofus.org). Summary statistics from the haematuria GWAS and PheWAS analyses are available upon reasonable request to the authors.

## Acknowledgements

This research was made possible through access to the data and findings generated by the UK Biobank and All of Us Research Program. The authors gratefully acknowledge the participation of all individuals recruited to these cohorts and their families. DPG is supported by the St Peter’s Trust for Kidney, Prostate and Bladder Research. KT is funded by KRUK studentship (ST_004_20221129); GTD is supported by KRUK fellowship (TF_001_20230627); OSA was funded by an MRC Clinical Research Training Fellowship (MR/S021329/1).

## Author Contributions

Design, conceptualisation and supervision: DPG; analysis: KT, GD, OSA. Each author contributed important intellectual content during manuscript drafting or revision.

