## Supplementary Figure for "Population-scale genomics reveals divergent pathogenicity of variant classes across paralogous collagen IV genes"

#### Supplemental Notes

##### Supplementary Note S1: *COL4A5* Per-Variant Association with Glomerular Haematuria

Given the X-linked inheritance of *COL4A5*, all analyses were performed separately in males and females. For haematuria (9,161 cases, 50.8% male; 335,746 controls, 42.6% male), 7 pathogenic coding variants were significantly associated in males and 12 in females (Figure S1a–b). Among these, glycine substitutions within the collagenous domain were the most frequent class in males (3 variants), while females showed a broader distribution including glycine substitutions (1 variant) and NC1 domain missense variants (3 variants). No truncating variants reached significance in either sex.

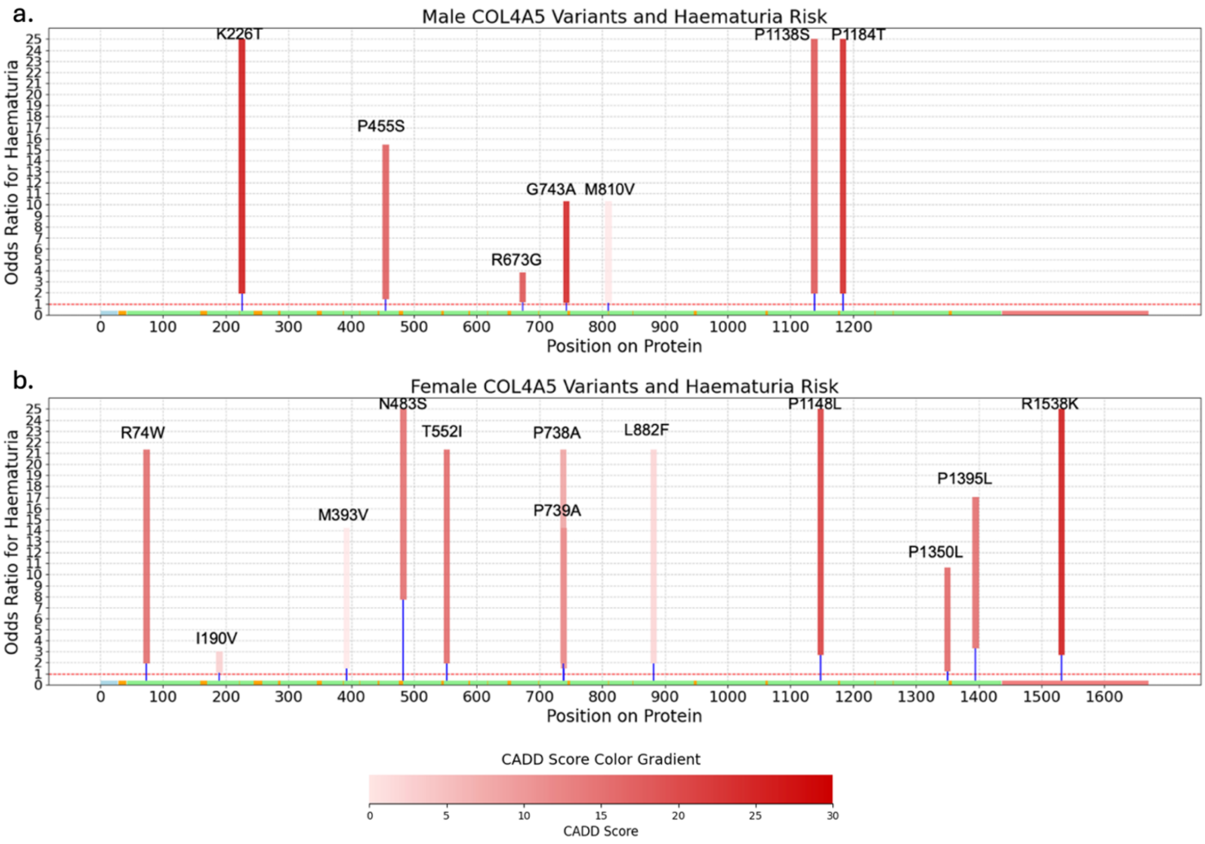


*Figure S1. COL4A5 Variants with 95% CIs Entirely above 1 for Glomerular Haematuria.*

*a. Males. b. Females. Protein-altering variants with 95% confidence intervals entirely above 1 are mapped to their respective protein domains. Rectangles represent individual variants; height reflects the OR range from the lower bound of the 95% CI to the point estimate. For visualisation purposes, ORs are capped at 25, variants exceeding this threshold are displayed at the cap. Variants are colour-coded by CADD score (pale pink to dark red gradient indicating increasing predicted pathogenicity; grey indicates CADD not available). Protein domains are indicated below: N-terminus (blue), collagenous domain (green), NC1 domain (red), and non-collagenous interruptions (orange). The dashed red line at OR=1 denotes the threshold for statistical significance.*

#### Supplementary Note S2: *COL4A5* Per-Variant Association with Glomerular Proteinuria

For proteinuria (16,074 cases, 41.8% male; 222,674 controls, 46.1% male), 5 pathogenic coding variants were significantly associated in males and 12 in females (Figure S2a–b). Glycine substitutions predominated in both sexes (males: 2 variants; females: 2 variants), with NC1 missense variants also observed (males: 1 variant; females: 1 variants).

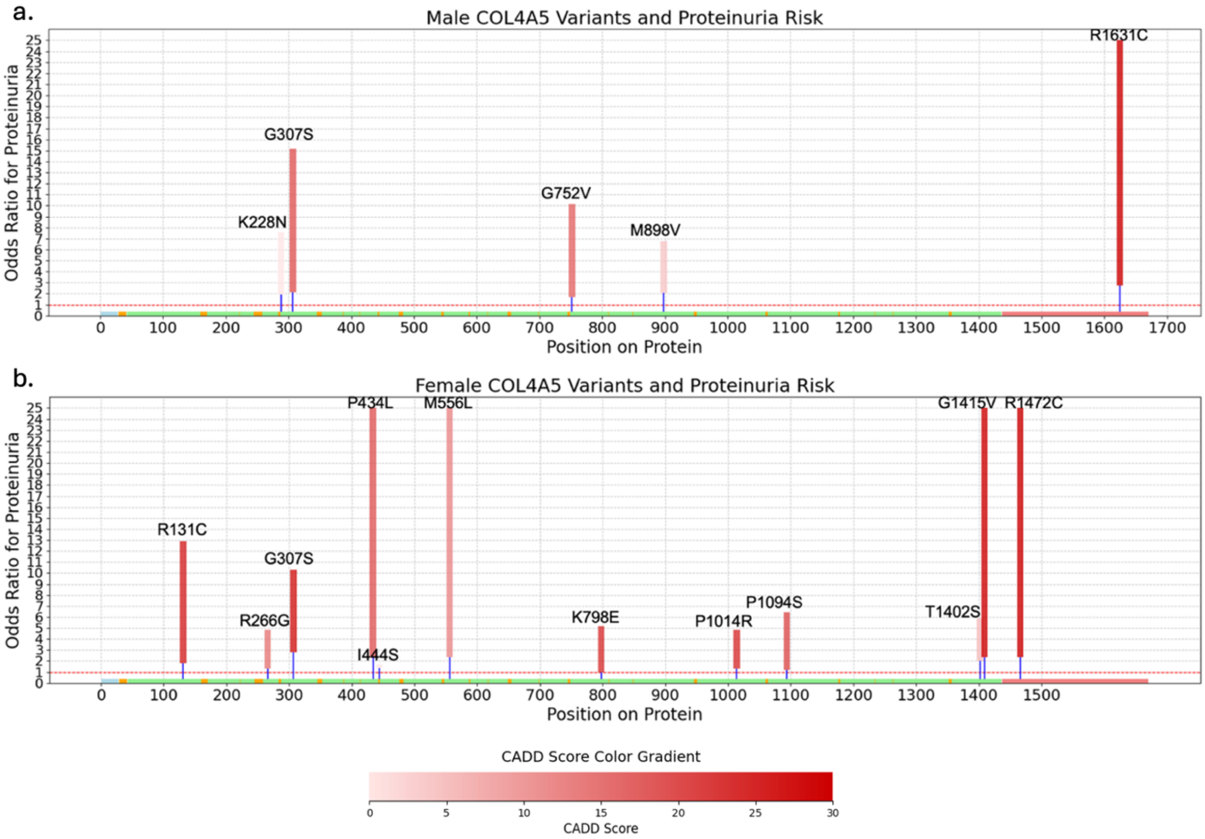


*Figure S2. COL4A5 Variants with 95% CIs Entirely above 1 for Glomerular Proteinuria.*

*a. Males. b. Females. As in Supplementary Figure 1, for proteinuria (uACR > 3 mg/mmol.*

#### Supplementary Note S3: *COL4A5* Per-Variant Association with Severe CKD

For severe CKD (4,335 cases, 61.7% male; 293,636 controls, 42.3% male), 9 pathogenic coding variants were significantly associated in males and 8 in females (Figure S3a–b). The distribution of variant classes was similar, with glycine substitutions (males: 3 variants; females: 1 variants) and NC1 missense variants (males: 0 variants; females: 3 variants) being the most frequent.

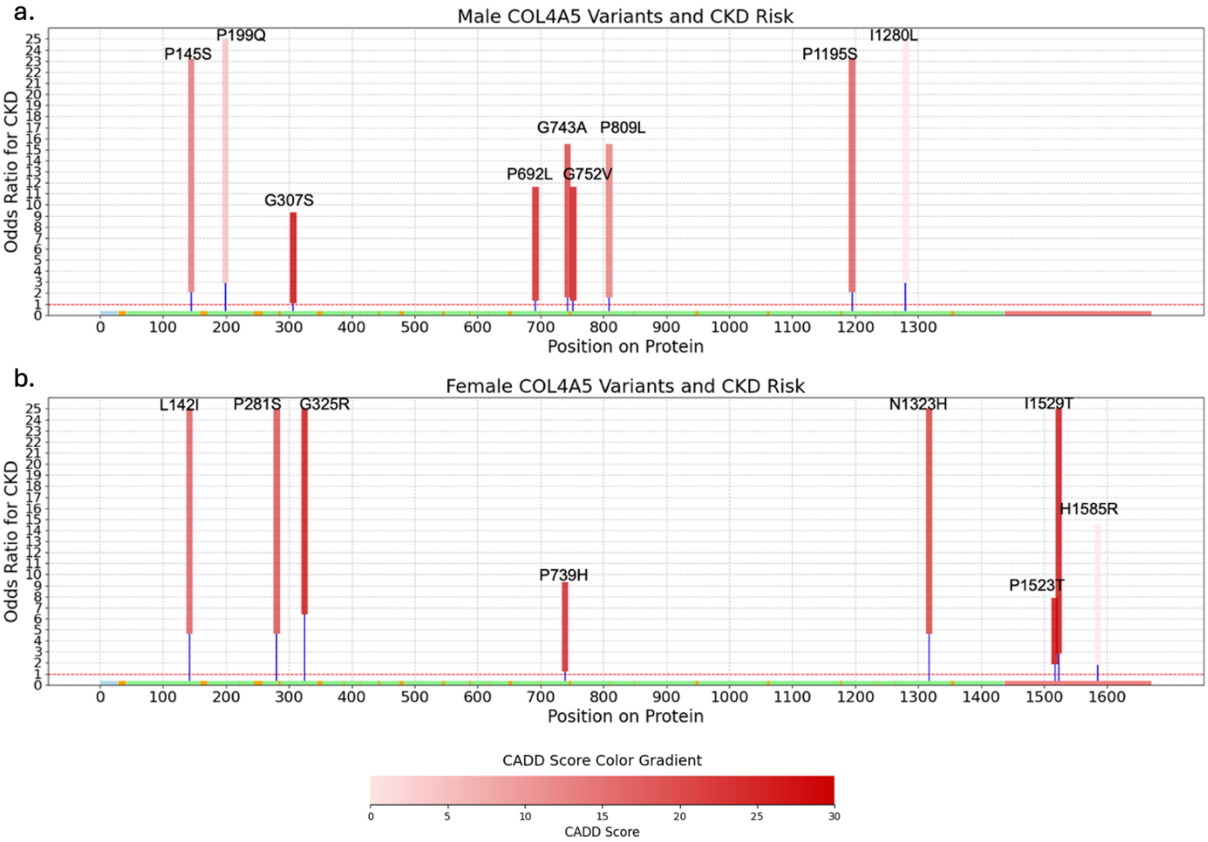


*Figure S3. COL4A5 Variants with 95% CIs Entirely above 1 for Severe CKD.*

*a. Males. b. Females. As in Supplementary Figure 1, for severe CKD (stage 4, stage 5 and end stage renal disease).*

#### Supplementary Note S4: CADD Score Distribution of NC1 Missense Variants in *COL4A3* and *COL4A4*

To assess whether differences in predicted pathogenicity might explain the gene-specific effects observed for NC1 domain missense variants, we compared the distribution of CADD scores between *COL4A3* and *COL4A4* variants included in the collapsing analyses (Figure S4). Both genes showed similar central tendencies, with *COL4A3* NC1 variants (n = 94) exhibiting a mean CADD score of 24.26 (median: 24.15) and *COL4A4* NC1 variants (n = 103) showing a mean of 23.98 (median: 24.20). The distributions substantially overlapped, with most variants in both genes falling within the CADD 20–25 range (*COL4A3*: 84.1%; *COL4A4*: 92.3%). These findings indicate that the stronger phenotypic effect observed for *COL4A4* NC1 variants cannot be attributed to higher predicted deleteriousness, as *COL4A3*

NC1 variants have comparable and indeed slightly higher CADD scores. This supports the interpretation that the gene-specific effect reflects inherent biological differences between *COL4A3* and *COL4A4* rather than differences in variant severity.

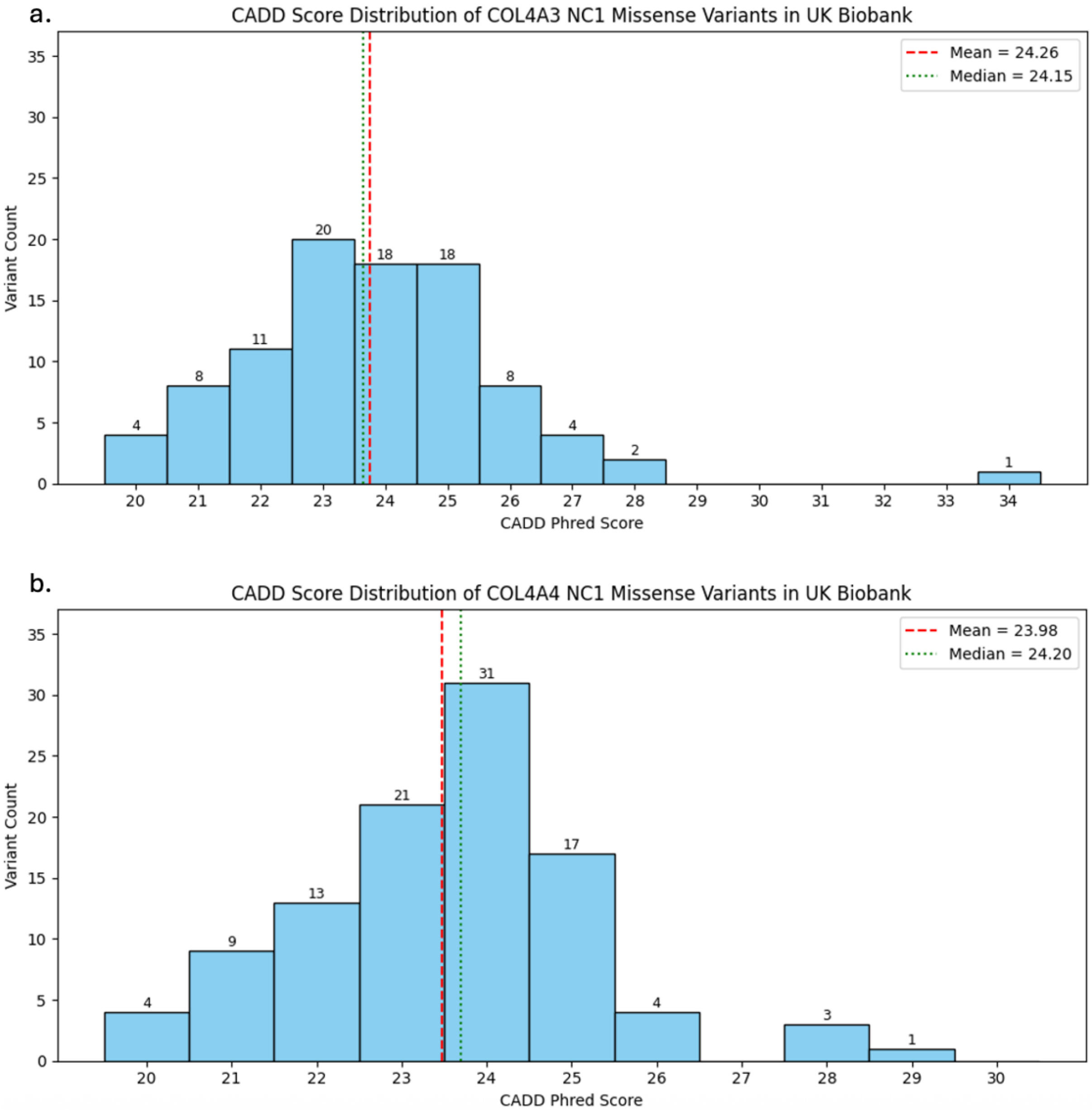


*Figure S4. Distribution of CADD scores for NC1 missense variants in COL4A3 and COL4A4.*

*Histograms showing the distribution of CADD scores for rare NC1 domain missense variants included in collapsing analyses. a. COL4A3 (n = 94 variants). b. COL4A4 (n = 103 variants). Blue vertical lines indicate mean CADD scores (COL4A3: 24.79; COL4A4: 24.27).*

#### Supplementary Note S5: REVEL Score Distribution of NC1 Missense Variants in *COL4A3* and *COL4A4*

To further assess predicted pathogenicity, we compared REVEL scores for NC1 missense variants in *COL4A3* and *COL4A4* (Figure S5). Both genes showed comparable distributions, with *COL4A3* variants (n = 94) having a mean REVEL score of 0.756 (median: 0.787; range: 0.214–0.982) and *COL4A4* variants (n = 103) a mean of 0.724 (median: 0.752; range: 0.209–0.962). The proportion of variants with REVEL ≥0.75 was similar between genes (*COL4A3*: 59.6%; *COL4A4*: 50.5%). These findings reinforce that the stronger phenotypic effect of *COL4A4* NC1 variants is not explained by differences in computational pathogenicity predictions.

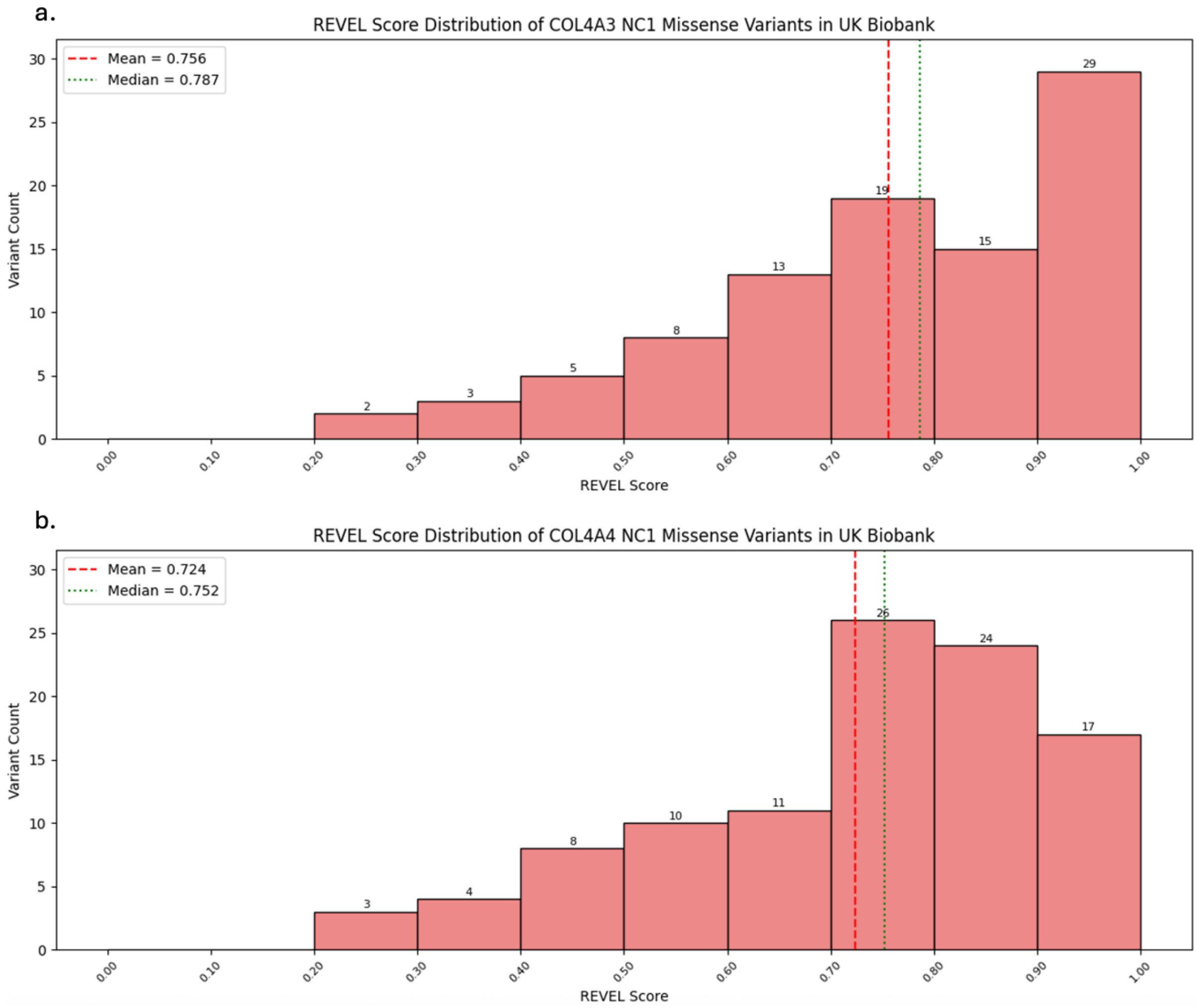


*Figure S5. Distribution of REVEL scores for NC1 missense variants in COL4A3 and COL4A4.*

*Histograms showing the distribution of REVEL scores for rare NC1 domain missense variants. a. COL4A3 (n = 102 variants). b. COL4A4 (n = 109 variants). Horizontal lines indicate median scores (COL4A3: 0.787; COL4A4: 0.752). Dashed red line at REVEL = 0.75 indicates the high-confidence pathogenicity threshold.*

#### Supplementary Note S6: Proteinuria Phenotype Definition in the All of Us Cohort

Unlike the UK Biobank, where urinary albumin and creatinine were measured uniformly at baseline assessment using standardized assays, the All of Us cohort comprises electronic health record data aggregated from multiple healthcare providers, resulting in substantial heterogeneity in laboratory measurements. Defining a binary proteinuria phenotype in this context required navigating several challenges.

First, measurements relevant to proteinuria were recorded under multiple distinct concept IDs, including microalbumin concentration, albumin-to-creatinine ratio (ACR), and urine dipstick results. For each measurement concept, data were recorded as either numeric values (with associated units) or categorical values (with predefined response options such as "negative", "trace", "+", "++", "+++"). Numeric measurements were recorded in a wide range of units, often inconsistently. For example, microalbumin concentration (concept: 3000034) was reported in at least 15 different units, including mg/dL, mg/L, µg/mL, and mg/g creatinine, alongside entries with no unit specified or with ambiguous descriptors such as "No matching concept" or “no value”. Categorical measurements, primarily from urine dipstick tests, contained a mix of ordinal descriptors and, in some cases, numeric entries with inconsistent units, further complicating harmonization. Additionally, there was no single laboratory test administered uniformly across the cohort; different individuals underwent different tests based on clinical indication, healthcare site and date, leading to missing data and variable ascertainment.

To address these challenges, we adopted a pragmatic approach. For numeric measurements, we first mapped all reported units to a common reference (mg/g creatinine for ACR, mg/dL for albumin concentration) using available conversion factors where possible. Measurements with no unit specified were examined by plotting their distribution; where a clear bimodal pattern emerged consistent with pathological versus normal ranges, values were retained and classified using phenotype-specific thresholds (e.g., uACR > 3 mg/mmol or equivalent). Measurements with units that could not be reliably converted or that fell outside plausible biological ranges were excluded. For categorical measurements, ordinal dipstick results were mapped to binary case/control status based on standard clinical cutoffs (e.g., "negative" or "trace" classified as control; "≥1+" classified as case). Measurements containing only non-informative entries (e.g., "No matching concept", "None") were excluded.

Despite these harmonization efforts, several limitations remain. The lack of a uniform measurement across the cohort means that proteinuria status could not be ascertained for all individuals, and misclassification, both false positives and false negatives, is possible. Individuals with proteinuria may have been missed if they lacked relevant laboratory tests, while individuals with borderline or transient proteinuria may have been incorrectly classified as cases. These limitations likely contribute to the attenuated replication signal for proteinuria in All of Us and should be considered when interpreting the results.

#### Supplementary Note S7: Collapsing PheWAS: Glycine Substitutions in All of Us

We replicated the collapsing PheWAS analyses in the All of Us Research Program cohort (v8), restricted to individuals of European ancestry aged ≥40 years (n ≈ 160,000), using identical variant classification and statistical methods (Bonferroni threshold $p < 3.8 \times 10^{-5}$). The six collapsing models were tested against 1,322 ICD-10 phenotypes. Glycine substitutions in both genes replicated the associations with haematuria observed in UK Biobank. *COL4A3* gly-sub carriers showed significant associations with unspecified haematuria (R31; $\beta = 0.025$, $p = 2.78 \times 10^{-6}$) and recurrent/persistent haematuria (N02; $\beta = 0.107$, $p = 2.94 \times 10^{-5}$) (Figure S6a). Proteinuria was the third strongest association ($\beta =$

0.041, p = 1.24 × $10^{-3}$) but did not reach significance. *COL4A4* gly-sub carriers were significantly associated with both R31 (β = 0.029, p = 4.34 × $10^{-14}$) and N02 (β = 0.112, p = 1.54 × $10^{-5}$), with proteinuria ranking third (β = 0.036, p = 2.89 × $10^{-3}$) (Figure S6b).

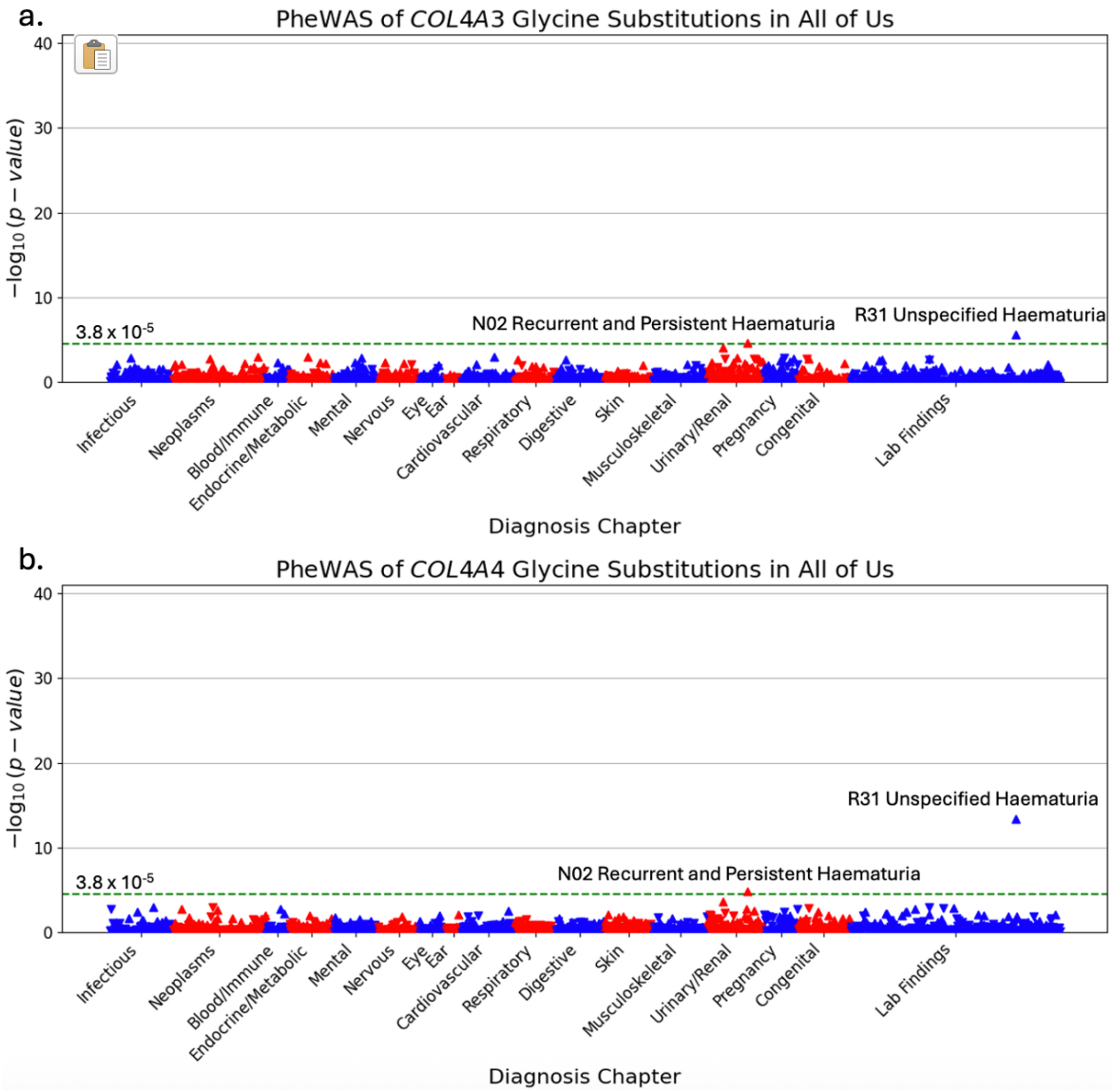


*Figure S6. Manhattan plots of diagnoses for collapsing tests of COL4A3/A4 glycine substitutions in AoU.*

*a. Manhattan plot showing the association of diagnoses with COL4A3 glycine substitutions. b. Manhattan plot diagnoses associated with glycine substitutions in COL4A4. Diagnoses are grouped by phenotypic categories on the x-axis. Upward-pointing triangles represent positive associations (OR > 1), while downward-pointing triangles represent negative associations (OR < 1). Red dashed line indicates the Bonferroni-corrected significance threshold.*

#### Supplementary Note S8: Collapsing PheWAS: Truncating Variants in All of Us

Truncating variants recapitulated the gene-specific pattern observed in UKB. *COL4A3* truncating variants showed no significant associations (strongest signal: R31, β = 0.001, p = 9.10 × $10^{-4}$; proteinuria: β = 0.019, p = 0.003) (Figure S7a). In contrast, *COL4A4* truncating variants were significantly associated with R31 (β = 0.064, p = 2.24 × $10^{-10}$) and unspecified kidney disorders (N28; β = 0.052, p = 1.11 × $10^{-5}$). Proteinuria ranked third (β = 0.067, p = 7.7 × $10^{-4}$) (Figure S7b).

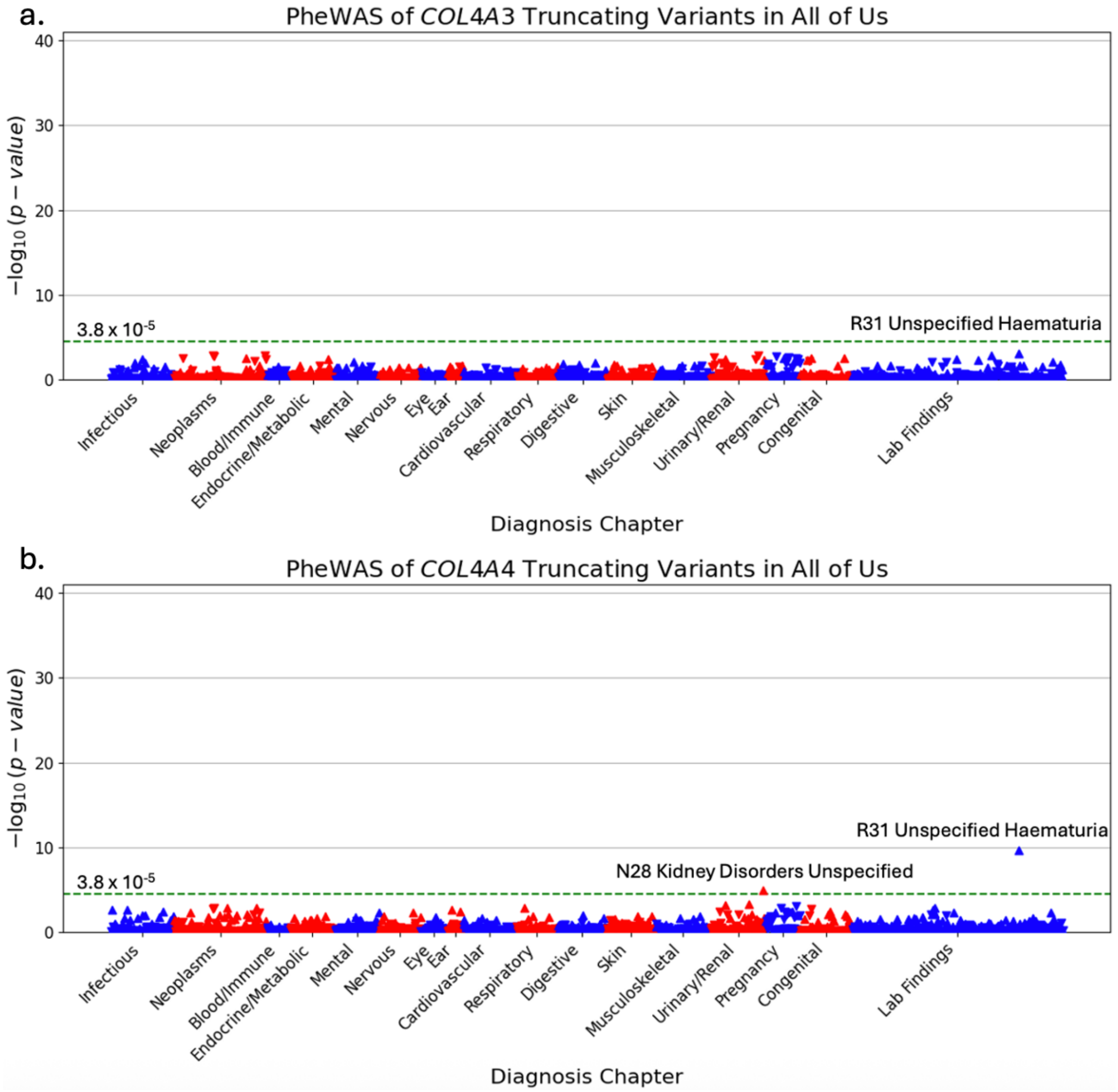


*Figure S7. Manhattan plots of diagnoses for collapsing tests of COL4A3/A4 truncating variants in AoU.*

*a. COL4A3 truncating variants. b. COL4A4 truncating variants. See Supplementary Figure 6 legend for detailed description of plot elements.*

#### Supplementary Note S9: Collapsing PheWAS: NC1 Missense Variants in All of Us

NC1 missense variants similarly showed no significant associations for COL4A3 (strongest signal: R31, $\beta = 0.005$, $p = 2.91 \times 10^{-4}$; proteinuria: $\beta = 0.005$, $p = 7.14 \times 10^{-4}$) (Figure S8a), while COL4A4 NC1 variants were significantly associated with R31 ($\beta = 0.024$, $p = 5.86 \times 10^{-7}$), with proteinuria ranked second for COL4A4 NC1 carriers ($\beta = 0.040$, $p = 4.24 \times 10^{-4}$) (Figure S8b).

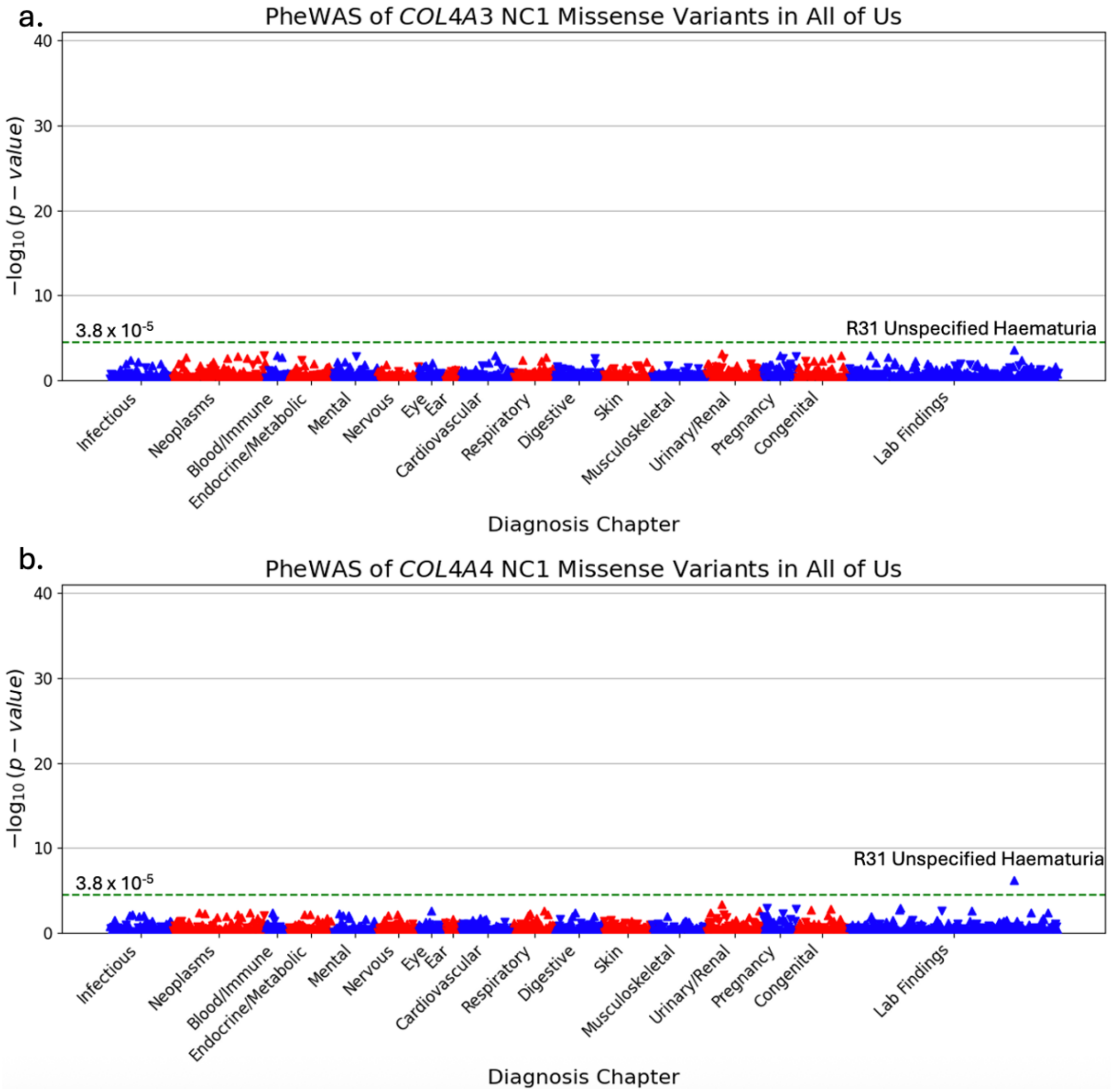


*Figure S8. Manhattan plots of diagnoses for collapsing tests of COL4A3/A4 NC1 domain missense variants in AoU.*

*a. COL4A3 NC1 missense variants. b. COL4A4 NC1 missense variants. See Supplementary Figure 6 legend for detailed description of plot elements.*

#### Supplementary Note S10: CADD Score Distribution of NC1 Missense Variants in *COL4A3* and *COL4A4* (All of Us)

To confirm that the gene-specific effects observed in All of Us were not driven by differences in predicted pathogenicity, we compared CADD score distributions for NC1 missense variants in the replication cohort (Figure S9). *COL4A3* NC1 variants (n = 74) had a mean CADD score of 24.68 (median: 24.55), with the majority falling within the CADD 20–25 range (81.1%). *COL4A4* NC1 variants (n = 97) showed a lower mean CADD score of approximately 23.9 (median: 24.1), with 94.7% of variants in the CADD 20–25 range. These findings mirror the UK Biobank results, confirming that the stronger phenotypic effect of *COL4A4* NC1 variants cannot be attributed to higher predicted deleteriousness, as *COL4A3* NC1 variants have comparable or greater CADD scores.

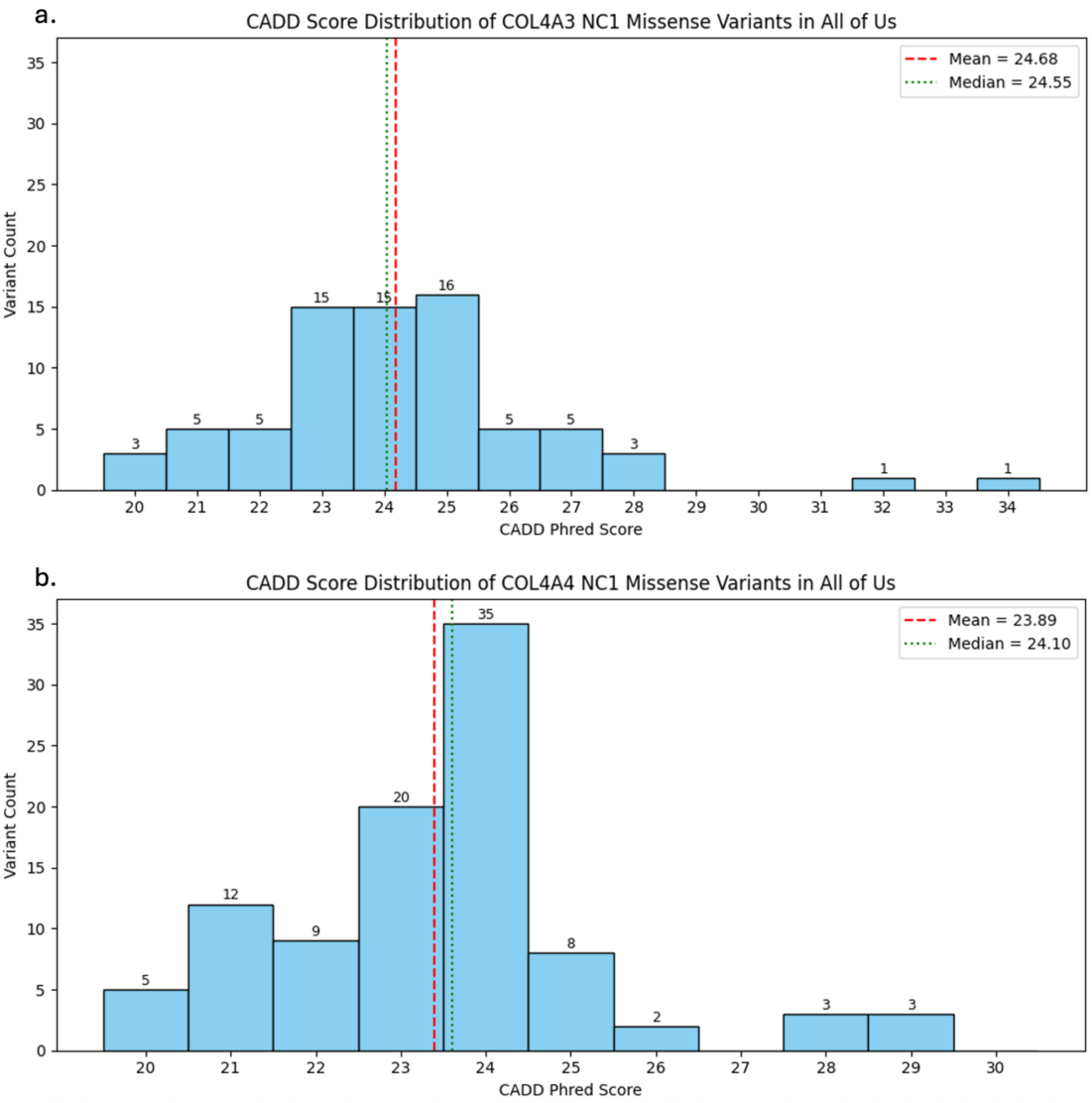


*Figure S9. Distribution of CADD scores for NC1 missense variants in COL4A3 and COL4A4 (All of Us).*

*Histograms showing the distribution of CADD scores for rare NC1 domain missense variants in the All of Us replication cohort. a. COL4A3 (n = 74 variants). b. COL4A4 (n = 97 variants). Blue vertical lines indicate mean CADD scores (COL4A3: 25.18; COL4A4: 24.27).*

#### Supplementary Note S11: REVEL Score Distribution of NC1 Missense Variants in *COL4A3* and *COL4A4* (All of Us)

To further assess predicted pathogenicity in the replication cohort, we compared REVEL scores for NC1 missense variants in *COL4A3* and *COL4A4* (Figure S10). *COL4A3* NC1 variants (n = 74) had a mean REVEL score of 0.796 (median: 0.851; range: 0.330–0.982), with 67.1% of variants scoring ≥0.75. *COL4A4* NC1 variants (n = 97) showed a lower mean REVEL score of 0.692 (median: 0.736; range: 0.156–0.975), with 45.4% of variants scoring ≥0.75. These findings reinforce that the stronger phenotypic effect observed for *COL4A4* NC1 variants in All of Us is not explained by higher computational pathogenicity predictions, as *COL4A3* NC1 variants demonstrate consistently higher REVEL scores across multiple thresholds.

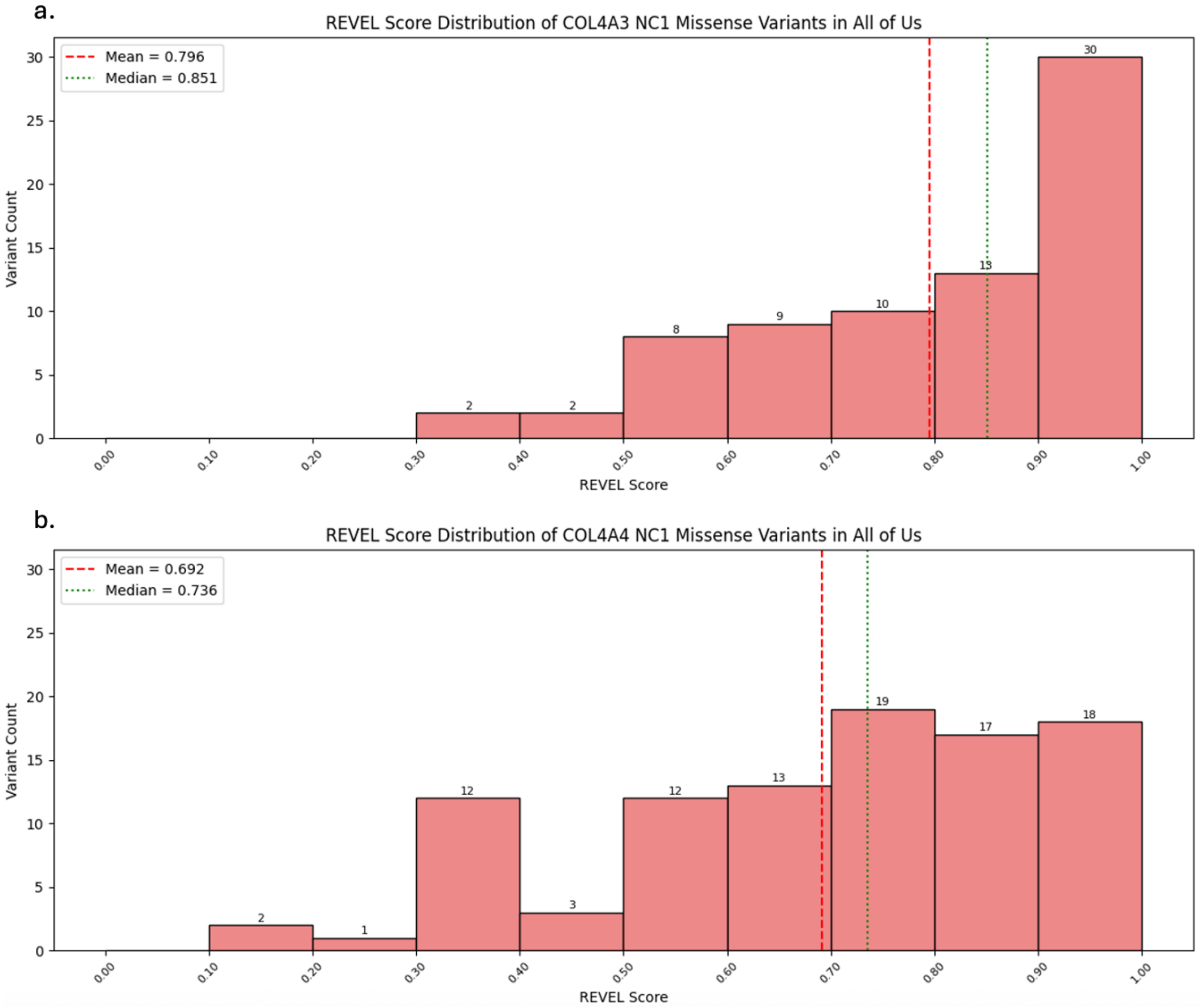


*Figure S10. Distribution of REVEL scores for NC1 missense variants in COL4A3 and COL4A4 (All of Us).*

*Histograms showing the distribution of REVEL scores for rare NC1 domain missense variants in the All of Us replication cohort. a. COL4A3 (n = 74 variants). b. COL4A4 (n = 97 variants). Horizontal lines indicate median scores (COL4A3: 0.851; COL4A4: 0.736). Dashed red line at REVEL = 0.75 indicates the high-confidence pathogenicity threshold.*

#### Supplementary Note S12: QQ Plots for Genome-Wide Association Studies of Haematuria

To assess potential genomic inflation and confirm that the GWAS results were not confounded by population stratification or technical artifacts, we generated quantile-quantile (QQ) plots for both the unadjusted and burden-adjusted GWAS (Figure S11). The unadjusted GWAS showed minimal inflation (λ GC = 1.029), with the observed p-values closely following the expected distribution under the null hypothesis except for the strong tail of significant associations at the *COL4A3/A4* locus. After adjustment for the weighted *COL4A3/A4* burden score (WBS), the inflation factor was further reduced (λ GC = 1.021), and the QQ plot showed no evidence of residual stratification or systematic bias. These findings confirm that the association signals identified are robust and that the adjustment for rare variant burden effectively controlled for the primary locus effect without introducing inflation.

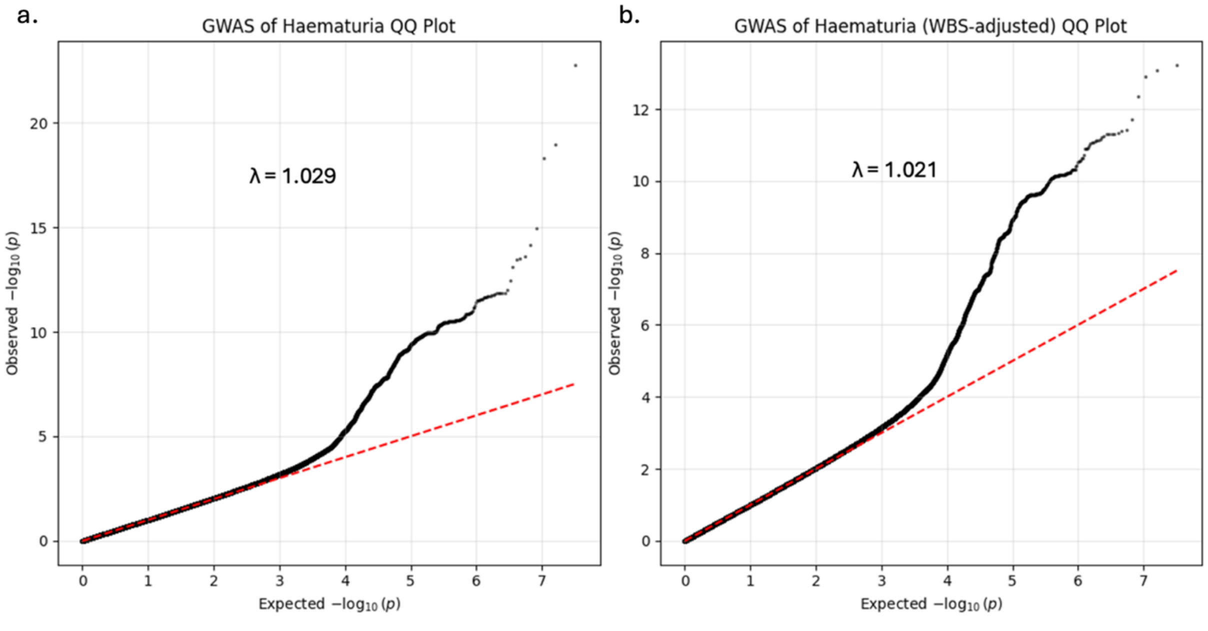


*Figure S11. Quantile-quantile (QQ) plots for haematuria GWAS.*

*a. Unadjusted GWAS. b. GWAS adjusted for weighted COL4A3/A4 burden score (WBS). Red lines indicate the expected distribution under the null hypothesis. Genomic inflation factors (λ GC) are shown in each panel.*

### Supplementary Note S13: Colocalization of the HLA-B Haematuria Signal with IgA Nephropathy

To characterise the *HLA-B* locus identified in the haematuria GWAS, we performed colocalization analysis using coloc.abf comparing the UKB haematuria GWAS summary statistics against a European IgA nephropathy meta-analysis (10,146 cases and 28,751 controls)[1]. The analysis was restricted to a ± 500 kb window around the haematuria lead SNP on chromosome 6 (crh6:31,474,954; rs2844503). The haematuria signal showed strong colocalization with the IgA nephropathy locus (PP.H4 = 0.989), with the posterior probability concentrated almost entirely on a single variant (6:31,869,050; rs521977).

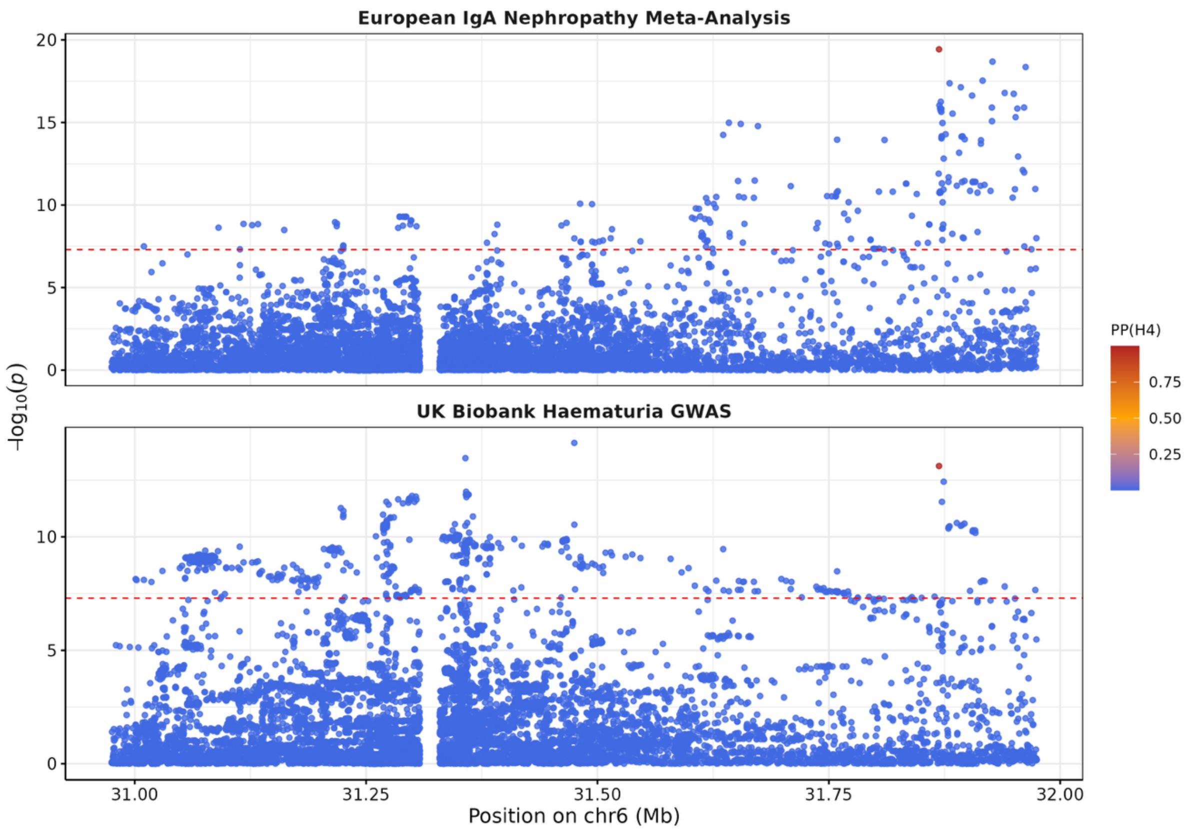


*Figure S12. Colocalization of the haematuria HLA-B signal with IgA Nephropathy.*

*Regional association plots for the HLA locus (chr6:30.97-31.97 Mb) in the European IgA Nephropathy Meta-Analysis (upper panel) and in the UK Biobank Haematuria GWAS (lower panel). Points are coloured by their posterior probability of sharing a causal variant with the haematuria signal (PP.H4). The dashed red line indicates the genome-wide significance threshold ($p = 5 \times 10^{-8}$).*

#### Supplementary Note S14: QQ Plots for GWAS of Haematuria restricted to *COL4A3/A4* Heterozygotes

To assess genomic inflation in the secondary GWAS restricted to heterozygotes for qualifying *COL4A3* and *COL4A4* variants, we generated QQ plots for both unadjusted and burden-adjusted analyses (Figure S13). The unadjusted GWAS in heterozygotes showed minimal inflation (λ GC = 1.031), with observed p-values closely following the expected distribution under the null hypothesis. The slight inflation to the primary GWAS (λ GC = 1.029) is consistent with the reduced sample size (1,627, 8,755 controls) rather than population stratification. After adjustment for the *COL4A3/A4* WBS, the inflation factor improved to λ GC = 1.021. Both QQ plots showed no evidence of systematic bias or residual inflation.

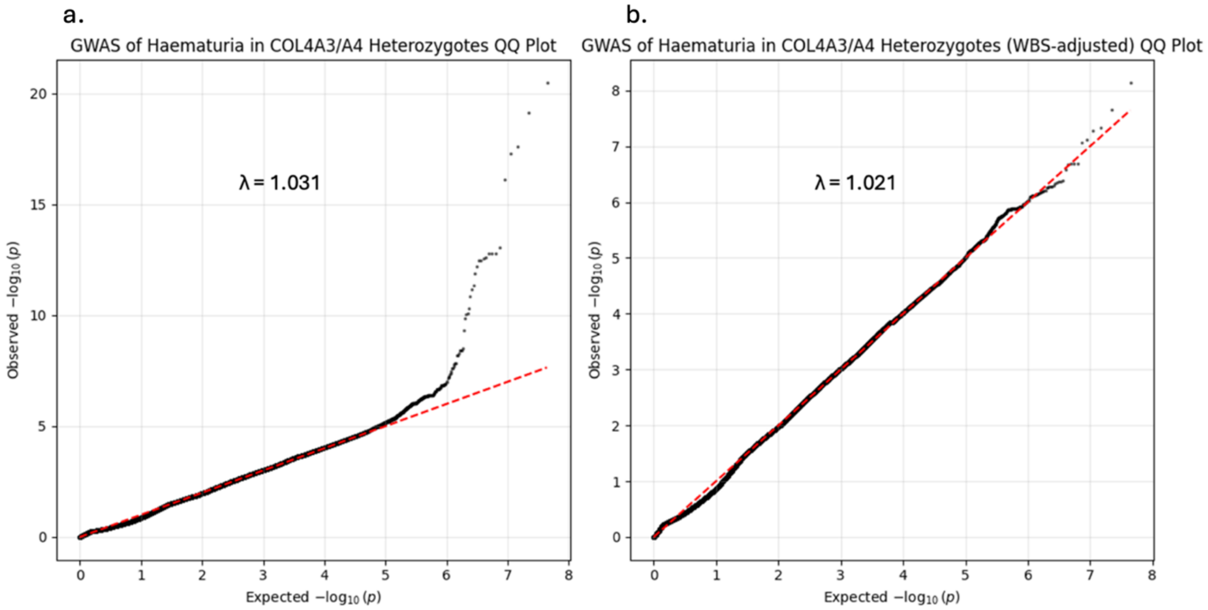


*Figure S13. Quantile-quantile (QQ) plots for haematuria GWAS restricted to COL4A3/A4 heterozygotes.*

*a. Unadjusted GWAS restricted to individuals carrying at least one qualifying COL4A3 or COL4A4 variant. b. GWAS in the same heterozygote-restricted population after adjustment for weighted COL4A3/A4 burden score (WBS). Red lines indicate the expected distribution under the null hypothesis. Genomic inflation factors (λ GC) are shown in each panel.*

#### Supplementary Note S15: Genome-Wide Association Study for Proteinuria

To identify genetic loci associated with proteinuria, we performed a GWAS of the urinary albumin-to-creatinine ratio (uACR) as a quantitative trait in 93,374 individuals from the UK Biobank. In total 19 distinct loci reached genome wide significant (Figure S14). The most strongly associated locus was at the CUBN gene on chromosome 10, which encodes cubulin, a proximal tubule endocytic receptor critical for albumin reabsorption, with the lead variant being the missense variant p.A1690V (rs141640975; $\beta = 0.03$; $p = 9.5 \times 10^{-54}$). The next most significant associated loci were *FRG1*, *COL4A3/A4* and *NPHS1* were the top four findings replicate those reported in the largest previously GWAS of uACR[2].

Several additional loci with prior evidence of association with albuminuria or uACR were identified, including *FOXD2*, *LRMDA*, *ACOXL*, *PRKCI*, *CCT2*, *AHR*, and *CYP1A1/A2*[2–4]. Additionally, *GCKR* and *DAB2* have previously been associated with eGFR and incident CKD [2,5]. Several further loci reached genome-wide significance but were supported by only one or two variants and had no prior evidence of association with kidney disease (Figure S14).

To determine whether the *COL4A3/A4* signal was driven by rare variant effects, we performed a second GWAS adjusting for the weighted *COL4A3/A4* WBS. The *COL4A3/A4* locus signal was completely abolished after adjustment, consistent with the haematuria findings and confirming that this signal is entirely attributable to the quantified rare variant effects. All remaining genome-wide significant loci were robust to WBS adjustment, and no new loci emerged, indicating no strong trans-acting modifiers of proteinuria risk in this population.

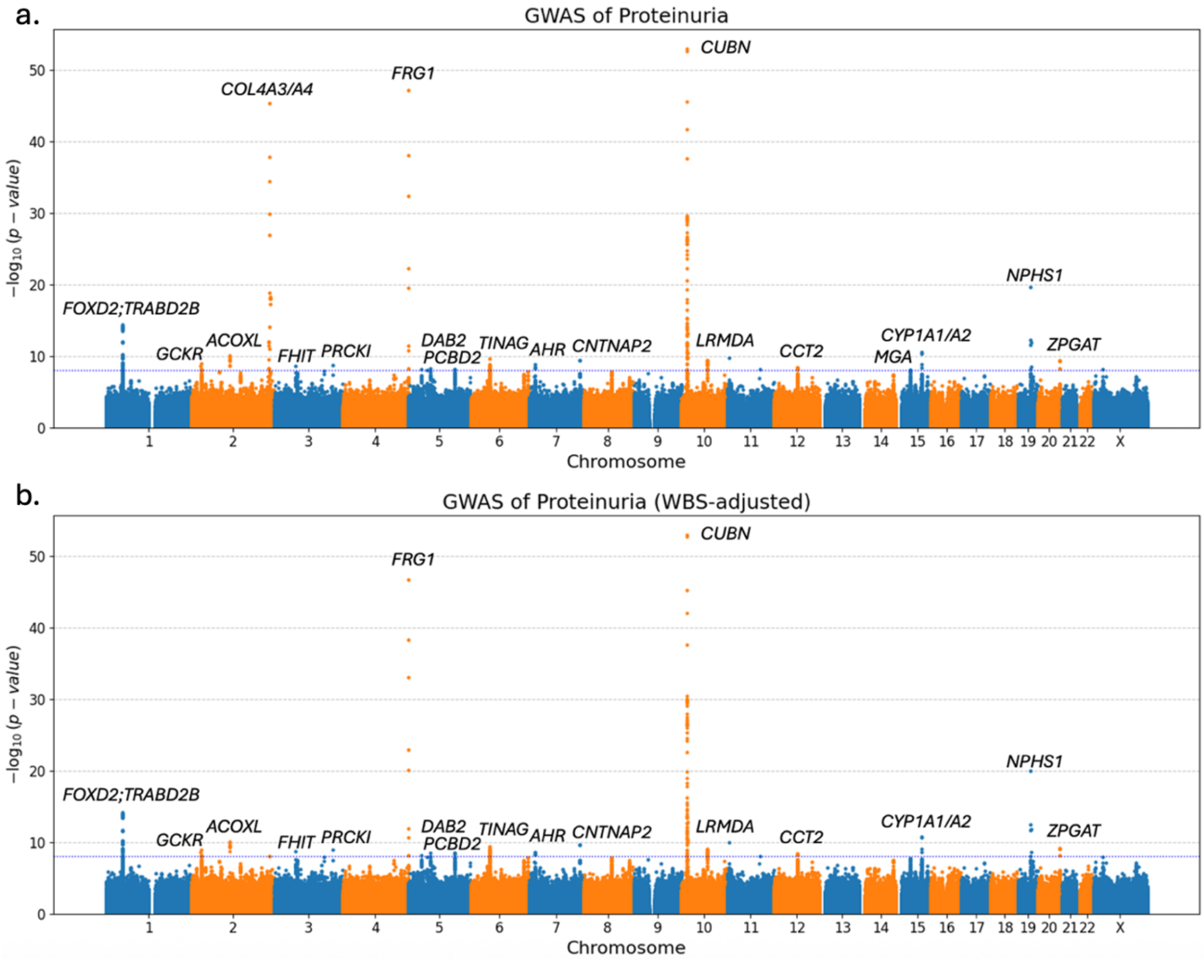


*Figure S14. Genome-wide association studies for proteinuria.*

*a. Manhattan plot showing association strength (-$\log_{10}$ P-value) across the genome for proteinuria (λ = 1.032). b. Manhattan plot after including WBS as a covariate (λ = 1.029). Red line indicates genome-wide significance threshold ($p < 5 \times 10^{-8}$).*

#### Supplementary Note S16: QQ Plots for Genome-Wide Association Studies of Proteinuria

To confirm that the GWAS results for proteinuria were not confounded by population stratification or technical artifacts, its genomic inflation was examined by generating QQ plots for both unadjusted and adjusted for WBS GWAS (Figure S15). The unadjusted GWAS showed minimal inflation (λ GC = 1.032), with observed p-values closely following the expected distribution under the null hypothesis, aside from the expected tail of significant associations. After adjustment for the *COL4A3/A4* WBS, the inflation factor was marginally improved (λ GC = 1.029) and the QQ plot showed no evidence of residual stratification or systematic bias. These findings confirm that the association signals for proteinuria are robust and that adjustment for rare *COL4A3/A4* variant burden effectively controlled for the *COL4A3/A4* locus without introducing inflation.

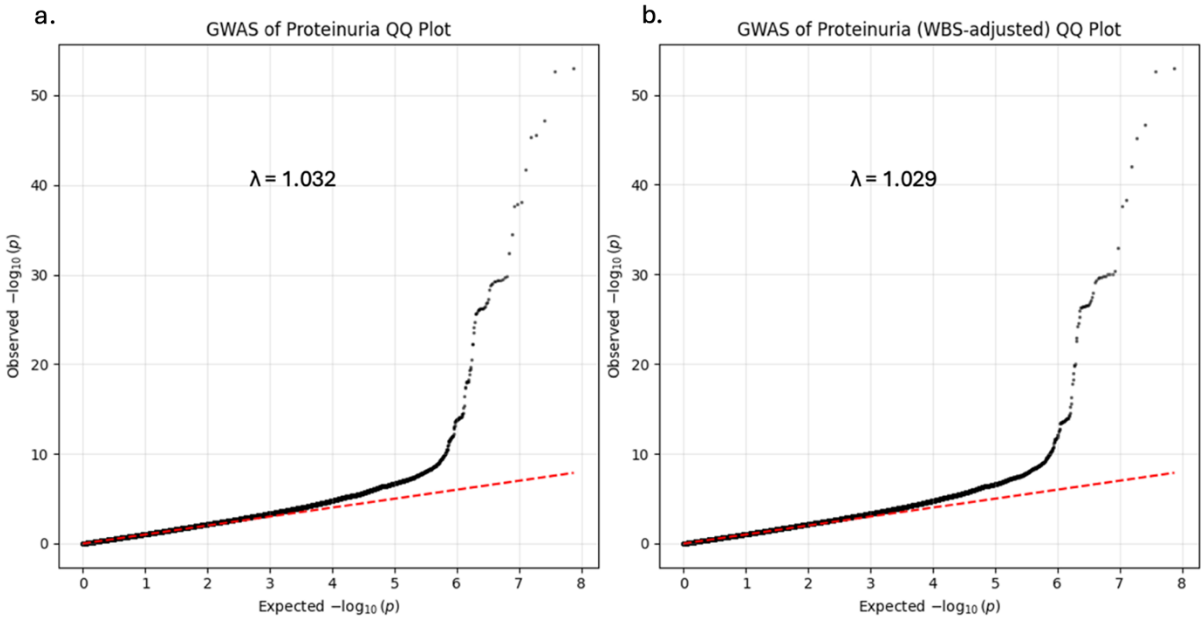


*Figure S15. Quantile-quantile (QQ) plots for proteinuria GWAS.*

*a. Unadjusted GWAS. b. GWAS adjusted for weighted COL4A3/A4 burden score (WBS). Red lines indicate the expected distribution under the null hypothesis. Genomic inflation factors (λ GC) are shown in each panel.*

### References

(1) Kiryluk, K.; Sanchez-Rodriguez, E.; Zhou, X.-J.; Zanoni, F.; Liu, L.; Mladkova, N.; Khan, A.; Marasa, M.; Zhang, J. Y.; Balderes, O.; Sanna-Cherchi, S.; Bomback, A. S.; Canetta, P. A.; Appel, G. B.; Radhakrishnan, J.; Trimarchi, H.; Sprangers, B.; Cattran, D. C.; Reich, H.; Pei, Y.; Ravani, P.; Galesic, K.; Maixnerova, D.; Tesar, V.; Stengel, B.; Metzger, M.; Canaud, G.; Maillard, N.; Berthoux, F.; Berthelot, L.; Pillebout, E.; Monteiro, R.; Nelson, R.; Wyatt, R. J.; Smoyer, W.; Mahan, J.; Samhar, A.-A.; Hidalgo, G.; Quiroga, A.; Weng, P.; Sreedharan, R.; Selewski, D.; Davis, K.; Kallash, M.; Vasylyeva, T. L.; Rheault, M.; Chishti, A.; Ranch, D.; Wenderfer, S. E.; Samsonov, D.; Claes, D. J.; Akchurin, O.; Goumenos, D.; Stangou, M.; Nagy, J.; Kovacs, T.; Fiaccadori, E.; Amoroso, A.; Barlassina, C.; Cusi, D.; Del Vecchio, L.; Battaglia, G. G.; Bodria, M.; Boer, E.; Bono, L.; Boscutti, G.; Caridi, G.; Lugani, F.; Ghiggeri, G.; Coppo, R.; Peruzzi, L.; Esposito, V.; Esposito, C.; Feriozzi, S.; Polci, R.; Frasca, G.; Galliani, M.; Garozzo, M.; Mitrotti, A.; Gesualdo, L.; Granata, S.; Zaza, G.; Londrino, F.; Magistroni, R.; Pisani, I.; Magnano, A.; Marcantoni, C.; Messa, P.; Mignani, R.; Pani, A.; Ponticelli, C.; Roccatello, D.; Salvadori, M.; Salvi, E.; Santoro, D.; Gembillo, G.; Savoldi, S.; Spotti, D.; Zamboli, P.; Izzi, C.; Alberici, F.; Delbarba, E.; Florczak, M.; Krata, N.; Mucha, K.; Pączek, L.; Niemczyk, S.;

Moszczuk, B.; Pańczyk-Tomaszewska, M.; Mizerska-Wasiak, M.; Perkowska-Ptasińska, A.; Bączkowska, T.; Durlik, M.; Pawlaczyk, K.; Sikora, P.; Zaniew, M.; Kaminska, D.; Krajewska, M.; Kuzmiuk-Glembin, I.; Heleniak, Z.; Bullo-Piontecka, B.; Liberek, T.; Dębska-Slizien, A.; Hryszko, T.; Materna-Kiryluk, A.; Miklaszewska, M.; Szczepańska, M.; Dyga, K.; Machura, E.; Siniewicz-Luzeńczyk, K.; Pawlak-Bratkowska, M.; Tkaczyk, M.; Runowski, D.; Kwella, N.; Drożdż, D.; Habura, I.; Kronenberg, F.; Prikhodina, L.; van Heel, D.; Fontaine, B.; Cotsapas, C.; Wijmenga, C.; Franke, A.; Annese, V.; Gregersen, P. K.; Parameswaran, S.; Weirauch, M.; Kottyan, L.; Harley, J. B.; Suzuki, H.; Narita, I.; Goto, S.; Lee, H.; Kim, D. K.; Kim, Y. S.; Park, J.-H.; Cho, B.; Choi, M.; Van Wijk, A.; Huerta, A.; Ars, E.; Ballarin, J.; Lundberg, S.; Vogt, B.; Mani, L.-Y.; Caliskan, Y.; Barratt, J.; Abeygunaratne, T.; Kalra, P. A.; Gale, D. P.; Panzer, U.; Rauen, T.; Floege, J.; Schlosser, P.; Ekici, A. B.; Eckardt, K.-U.; Chen, N.; Xie, J.; Lifton, R. P.; Loos, R. J. F.; Kenny, E. E.; Ionita-Laza, I.; Köttgen, A.; Julian, B. A.; Novak, J.; Scolari, F.; Zhang, H.; Gharavi, A. G. Genome-Wide Association Analyses Define Pathogenic Signaling Pathways and Prioritize Drug Targets for IgA Nephropathy. *Nat Genet* **2023**, *55* (7), 1091–1105. https://doi.org/10.1038/s41588-023-01422-x.

(2) Teumer, A.; Li, Y.; Ghasemi, S.; Prins, B. P.; Wuttke, M.; Hermle, T.; Giri, A.; Sieber, K. B.; Qiu, C.; Kirsten, H.; Tin, A.; Chu, A. Y.; Bansal, N.; Feitosa, M. F.; Wang, L.; Chai, J.-F.; Cocca, M.; Fuchsberger, C.; Gorski, M.; Hoppmann, A.; Horn, K.; Li, M.; Marten, J.; Noce, D.; Nutile, T.; Sedaghat, S.; Sveinbjornsson, G.; Tayo, B. O.; van der Most, P. J.; Xu, Y.; Yu, Z.; Gerstner, L.; Ärnlöv, J.; Bakker, S. J. L.; Baptista, D.; Biggs, M. L.; Boerwinkle, E.; Brenner, H.; Burkhardt, R.; Carroll, R. J.; Chee, M.-L.; Chee, M.-L.; Chen, M.; Cheng, C.-Y.; Cook, J. P.; Coresh, J.; Corre, T.; Danesh, J.; de Borst, M. H.; De Grandi, A.; de Mutsert, R.; de Vries, A. P. J.; Degenhardt, F.; Dittrich, K.; Divers, J.; Eckardt, K.-U.; Ehret, G.; Endlich, K.; Felix, J. F.; Franco, O. H.; Franke, A.; Freedman, B. I.; Freitag-Wolf, S.; Gansevoort, R. T.; Giedraitis, V.; Gögele, M.; Grundner-Culemann, F.; Gudbjartsson, D. F.; Gudnason, V.; Hamet, P.; Harris, T. B.; Hicks, A. A.; Holm, H.; Foo, V. H. X.; Hwang, S.-J.; Ikram, M. A.; Ingelsson, E.; Jaddoe, V. W. V.; Jakobsdottir, J.; Josyula, N. S.; Jung, B.; Kähönen, M.; Khor, C.-C.; Kiess, W.; Koenig, W.; Körner, A.; Kovacs, P.; Kramer, H.; Krämer, B. K.; Kronenberg, F.; Lange, L. A.; Langefeld, C. D.; Lee, J. J.-M.; Lehtimäki, T.; Lieb, W.; Lim, S.-C.; Lind, L.; Lindgren, C. M.; Liu, J.; Loeffler, M.; Lyytikäinen, L.-P.; Mahajan, A.; Maranville, J. C.; Mascalzoni, D.; McMullen, B.; Meisinger, C.; Meitinger, T.; Miliku, K.; Mook-Kanamori, D. O.; Müller-Nurasyid, M.; Mychaleckyj, J. C.; Nauck, M.; Nikus, K.; Ning, B.; Noordam, R.; Connell, J. O.; Olafsson, I.; Palmer, N. D.; Peters, A.; Podgornaia, A. I.; Ponte, B.; Poulain, T.; Pramstaller, P. P.; Rabelink, T. J.; Raffield, L. M.; Reilly, D. F.; Rettig, R.; Rheinberger, M.; Rice, K. M.; Rivadeneira, F.; Runz, H.; Ryan, K. A.; Sabanayagam, C.; Saum, K.-U.; Schöttker, B.; Shaffer, C. M.; Shi, Y.; Smith, A. V.; Strauch, K.; Stumvoll, M.; Sun, B. B.; Szymczak, S.; Tai, E.-S.; Tan, N. Y. Q.; Taylor, K. D.; Teren, A.; Tham, Y.-C.; Thiery, J.; Thio, C. H. L.; Thomsen, H.; Thorsteinsdottir, U.; Tönjes, A.; Tremblay, J.; Uitterlinden, A. G.; van der Harst, P.; Verweij, N.; Vogelezang, S.; Völker, U.; Waldenberger, M.; Wang, C.; Wilson, O. D.; Wong, C.; Wong, T.-Y.; Yang, Q.; Yasuda, M.; Akilesh, S.; Bochud, M.; Böger, C. A.; Devuyst, O.; Edwards, T. L.; Ho, K.; Morris, A. P.; Parsa, A.; Pendergrass, S. A.; Psaty, B. M.; Rotter, J. I.; Stefansson, K.; Wilson, J. G.; Susztak, K.; Snieder, H.; Heid, I. M.; Scholz, M.; Butterworth, A. S.; Hung, A. M.; Pattaro, C.; Köttgen, A. Genome-Wide Association Meta-Analyses and Fine-Mapping Elucidate Pathways Influencing Albuminuria. *Nat Commun* **2019**, *10*, 4130. https://doi.org/10.1038/s41467-019-11576-0.

(3) Haas, M. E.; Aragam, K. G.; Emdin, C. A.; Bick, A. G.; Hemani, G.; Davey Smith, G.; Kathiresan, S. Genetic Association of Albuminuria with Cardiometabolic Disease and Blood Pressure. *Am J Hum Genet* **2018**, *103* (4), 461–473. https://doi.org/10.1016/j.ajhg.2018.08.004.

(4) Verma, A.; Huffman, J. E.; Rodriguez, A.; Conery, M.; Liu, M.; Ho, Y.-L.; Kim, Y.; Heise, D. A.; Guare, L.; Panickan, V. A.; Garcon, H.; Linares, F.; Costa, L.; Goethert, I.; Tipton, R.; Honerlaw, J.; Davies, L.; Whitbourne, S.; Cohen, J.; Posner, D. C.; Sangar, R.; Murray, M.; Wang, X.; Dochtermann, D. R.; Devineni, P.; Shi, Y.; Nandi, T. N.; Assimes, T.

L.; Brunette, C. A.; Carroll, R. J.; Clifford, R.; Duvall, S.; Gelernter, J.; Hung, A.; Iyengar, S. K.; Joseph, J.; Kember, R.; Kranzler, H.; Kripke, C. M.; Levey, D.; Luoh, S.-W.; Merritt, V. C.; Overstreet, C.; Deak, J. D.; Grant, S. F. A.; Polimanti, R.; Roussos, P.; Shakt, G.; Sun, Y. V.; Tsao, N.; Venkatesh, S.; Voloudakis, G.; Justice, A.; Begoli, E.; Ramoni, R.; Tourassi, G.; Pyarajan, S.; Tsao, P.; O'Donnell, C. J.; Muralidhar, S.; Moser, J.; Casas, J. P.; Bick, A. G.; Zhou, W.; Cai, T.; Voight, B. F.; Cho, K.; Gaziano, J. M.; Madduri, R. K.; Damrauer, S.; Liao, K. P. Diversity and Scale: Genetic Architecture of 2068 Traits in the VA Million Veteran Program. *Science* **2024**, *385* (6706), eadj1182. https://doi.org/10.1126/science.adj1182.
(5) Wuttke, M.; Li, Y.; Li, M.; Sieber, K. B.; Feitosa, M. F.; Gorski, M.; Tin, A.; Wang, L.; Chu, A. Y.; Hoppmann, A.; Kirsten, H.; Giri, A.; Chai, J.-F.; Sveinbjornsson, G.; Tayo, B. O.; Nutile, T.; Fuchsberger, C.; Marten, J.; Cocca, M.; Ghasemi, S.; Xu, Y.; Horn, K.; Noce, D.; van der Most, P. J.; Sedaghat, S.; Yu, Z.; Akiyama, M.; Afaq, S.; Ahluwalia, T. S.; Almgren, P.; Amin, N.; Ärnlöv, J.; Bakker, S. J. L.; Bansal, N.; Baptista, D.; Bergmann, S.; Biggs, M. L.; Biino, G.; Boehnke, M.; Boerwinkle, E.; Boissel, M.; Bottinger, E. P.; Boutin, T. S.; Brenner, H.; Brumat, M.; Burkhardt, R.; Butterworth, A. S.; Campana, E.; Campbell, A.; Campbell, H.; Canouil, M.; Carroll, R. J.; Catamo, E.; Chambers, J. C.; Chee, M.-L.; Chee, M.-L.; Chen, X.; Cheng, C.-Y.; Cheng, Y.; Christensen, K.; Cifkova, R.; Ciullo, M.; Pina Concas, M.; Cook, J. P.; Coresh, J.; Corre, T.; Sala, C. F.; Cusi, D.; Danesh, J.; Daw, E. W.; de Borst, M. H.; De Grandi, A.; de Mutsert, R.; de Vries, A. P. J.; Degenhardt, F.; Delgado, G.; Demirkan, A.; Di Angelantonio, E.; Dittrich, K.; Divers, J.; Dorajoo, R.; Eckardt, K.-U.; Ehret, G.; Elliott, P.; Endlich, K.; Evans, M. K.; Felix, J. F.; Foo, V. H. X.; Franco, O. H.; Franke, A.; Freedman, B. I.; Freitag-Wolf, S.; Friedlander, Y.; Froguel, P.; Gansevoort, R. T.; Gao, H.; Gasparini, P.; Gaziano, J. M.; Giedraitis, V.; Gieger, C.; Girotto, G.; Giulianini, F.; Gögele, M.; Gordon, S. D.; Gudbjartsson, D. F.; Gudnason, V.; Haller, T.; Hamet, P.; Harris, T. B.; Hartman, C. A.; Hayward, C.; Hellwege, J. N.; Heng, C.-K.; Hickst, A. A.; Hofer, E.; Huang, W.; Hutri-Kähönen, N.; Hwang, S.-J.; ikram, M. A.; indridason, O. S.; Ingelsson, E.; ising, M.; Jaddoe, V. W. V.; Jakobsdottir, J.; Jonas, J. B.; Joshi, P. K.; Shilpa Josyula, N.; Jung, B.; Kähönen, M.; Kamatani, Y.; Kammerer, C. M.; Kanai, M.; Kastarinen, M.; Kerr, S. M.; Khor, C.-C.; Kiess, W.; Kleber, M. E.; Koenig, W.; Kooner, J. S.; Körner, A.; Kovacs, P.; Kraja, A. T.; Krajcoviechova, A.; Kramer, H.; Krämer, B. K.; Kronenberg, F.; Kubo, M.; Kühnel, B.; Kuokkanen, M.; Kuusisto, J.; La Bianca, M.; Laakso, M.; Lange, L. A.; Langefeld, C. D.; Jen-Mai Lee, J.; Lehne, B.; Lehtimäki, T.; Lieb, W.; Cohort Study, L.; Lim, S.-C.; Lind, L.; Lindgren, C. M.; Liu, J.; Liu, J.; Loeffler, M.; Loos, R. J. F.; Lucae, S.; Ann Lukas, M.; Lyytikäinen, L.-P.; Mägi, R.; Magnusson, P. K. E.; Mahajan, A.; Martin, N. G.; Martins, J.; März, W.; Mascalzoni, D.; Matsuda, K.; Christa Meisinger; Meitinger, T.; Melander, O.; Metspalu, A.; Mikaelsdottir, E. K.; Milaneschi, Y.; Miliku, K.; Mishra, P. P.; Veteran Program, V. A. M.; Mohlke, K. L.; Mononen, N.; Montgomery, G. W.; Mook-Kanamori, D. O.; Mychaleckyj, J. C.; Nadkarni, G. N.; Nalls, M. A.; Nauck, M.; Nikus, K.; Ning, B.; Nolte, ilja M.; Noordam, R.; O'Connell, J.; O'Donoghue, M. L.; Olafsson, I.; Oldehinkel, A. J.; Orho-Melander, M.; Ouwehand, W. H.; Padmanabhan, S.; Palmer, N. D.; Palsson, R.; Penninx, B. W. J. H.; Perls, T.; Perola, M.; Pirastu, M.; Pirastu, N.; Pistis, G.; Podgornaia, A. I.; Polasek, O.; Ponte, B.; Porteous, D. J.; Poulain, T.; Pramstaller, P. P.; Preuss, M. H.; Prins, B. P.; Province, M. A.; Rabelink, T. J.; Raffield, L. M.; Raitakari, O. T.; Reilly, D. F.; Rettig, R.; Rheinberger, M.; Rice, K. M.; Ridker, P. M.; Rivadeneira, F.; Rizzi, F.; Roberts, D. J.; Robino, A.; Rossing, P.; Rudan, I.; Rueedi, R.; Ruggiero, D.; Ryan, K. A.; Saba, Y.; Sabanayagam, C.; Salomaa, V.; Salvi, E.; Saum, K.-U.; Schmidt, H.; Schmidt, R.; Schöttker, B.; Schulz, C.-A.; Schupf, N.; Shaffer, C. M.; Shi, Y.; Smith, A. V.; Smith, B. H.; Soranzo, N.; Spracklen, C. N.; Strauch, K.; Stringham, H. M.; Stumvoll, M.; Svensson, P. O.; Szymczak, S.; Tai, E.-S.; Tajuddin, S. M.; Tan, N. Y. Q.; Taylor, K. D.; Teren, A.; Tham, Y.-C.; Thiery, J.; Thio, C. H. L.; Thomsen, H.; Thorleifsson, G.; Toniolo, D.; Tönjes, A.; Tremblay, J.; Tzoulaki, I.; Uitterlinden, A. G.; Vaccargiu, S.; van Dam, R. M.; van der Harst, P.; van Duijn, C. M.; Velez Edward, D. R.; Verweij, N.; Vogelezang, suzanne; Völker, üwe; Vollenweider, P.; Waeber, G.; Waldenberger, M.; Wallentin, L.;

Wang, Y. X.; Wang, C.; Waterworth, D. M.; Bin Wei, W.; White, H.; Whitfield, J. B.; Wild, S. H.; Wilson, J. F.; Wojczynski, M. K.; Wong, C.; Wong, T.-Y.; Xu, L.; Yang, Q.; Yasuda, M.; Yerges-Armstrong, L. M.; Zhang, W.; Zonderman, A. B.; Rotter, J. I.; Bochud, M.; Psaty, B. M.; Vitart, V.; Wilson, J. G.; Dehghan, A.; Parsa, A.; Chasman, D. I.; Ho, K.; Morris, A. P.; Devuyst, O.; Akilesh, S.; Pendergrass, S. A.; Sim, X.; Böger, C. A.; Okada, Y.; Edwards, T. L.; Snieder, H.; Stefansson, K.; Hung, A. M.; Heid, I. M.; Markus Scholz; Teumer, A.; Köttgen, A.; Pattaro, C. A Catalog of Genetic Loci Associated with Kidney Function from Analyses of a Million Individuals. *Nat Genet* **2019**, *51* (6), 957–972. https://doi.org/10.1038/s41588-019-0407-x.
